# Factors supporting the performance of primary care physician practices in Benin: a multiple case study

**DOI:** 10.1101/2024.03.08.24303725

**Authors:** Kéfilath Bello, Jan De Lepeleire, Djimon Marcel Zannou, Bart Criel

## Abstract

**Introduction:** In Benin, as in many African countries, the number of primary care physicians (PCPs) is gradually increasing. The literature suggests that these PCPs hold significant potential to improve the quality of care. However, this potential is not automatically realised. This study aims to understand the factors that underpin the performance of PCP practices in Benin.

**Methods:** We conducted a multiple case study of eight PCP practices with contrasting performance across five health districts in Benin. Both quantitative and qualitative methods were used. Quantitative data from consultation observations were used to assign performance scores, while qualitative data, gathered through 40 interviews and 16 focus group discussions with diverse stakeholders, underwent thematic analysis to identify influencing factors. A cross-case analysis was then used to determine the key factors supporting the performance of PCP practices in Benin.

**Results:** PCP performance scores varied, with communication scores ranging from 14.7 to 19.3 (out of 20) and technical quality scores from 68% to 88% (out of 100%). The overall performance of the PCP practices also showed substantial variation. The study identified various potential performance factors through key informant inputs and cross-case analysis. Among these, nine emerged from the cross-case analysis as factors supporting the performance of PCP practices: (i) values guiding PCP practices, (ii) preparation of PCPs for first-line practice, (iii) support from hierarchy, peers, or professional associations, (iv) leadership mandate and autonomy, (v) financing modalities, (vi) accountability mechanisms, (vii) PCPs’ relationship with their primary care team and their leadership style, (viii) collaboration with community leaders and public officials, and (ix) the contextual setting of each PCP practice. The study also examined causal pathways linking these factors to the performance of the PCPs’ practices.

**Conclusion:** This study has identified a series of factors that could serve as levers to enhance PCP practices in Benin.

## Introduction

Like the rest of the international community, African governments have committed to ensuring healthy lives and promoting well-being for all citizens [1]. Primary care, defined as the provision of first-contact care that is universally accessible, person-centred, continuous, comprehensive, and coordinated, delivered by a team of professionals accountable for addressing a large majority of the population’s healthcare needs for a given community, in a sustained partnership with this community [2], is proven to contribute to this goal [3].

In many African countries, primary care is offered in first-line facilities (also called primary care facilities), which serve as the first level of care or the entry point into the healthcare system at the interface between services and the community [4]. A high-quality primary care cannot be obtained without well-trained, motivated, available, and properly distributed human resources for health [5]. While the shortage of health workers is the most visible and debated issue when it comes to human resources for health in Africa [5], the sub-optimal utilisation and performance of the existing workforce is a significant, yet often unnoticed, problem.

Notably, in recent years, several African countries have seen a growing presence of primary care physicians (PCPs) [6,7], physicians working at primary care facilities and providing all-around care to the population, without distinction based on age, sex, or clinical condition [7]. This increase contrasts with the dominating policy choice which favours task shifting and provision of primary care services by nurse practitioners [8,9]. This increase results from a combination of physician training efforts by African universities, governments’ limited capacity to recruit and position them at higher healthcare levels, limited access to specialised training, and expansion of the private sector [10].

This growing presence of PCPs in Africa is mainly observed in the private sector (and urban areas) as a spontaneous, opportunistic, and poorly planned movement. However, there are a few cases where the PCPs are positioned as part of the academic development of family medicine or as part of a governmental choice to improve health systems’ outcomes [7]. For example, with its latest health policy (2018-2030), Benin has started to post physicians in some of the largest primary care facilities that were previously staffed by non-physicians. The aim is, among others, to improve the quality of care and financial viability of these facilities [11].

Several scholars have indicated that PCPs have a good potential for improving the quality of primary care in sub-Saharan Africa and the population’s well-being [7,12–14]. In many settings, PCPs improved the technical quality of care [15,16], the access to care for complex conditions [15,17,18], and the clinical processes through training and supervision of non-physicians [17]. They were also able to innovate during the COVID-19 pandemic and support the countries’ responses [12,14]. However, there is also evidence that PCPs in African countries can sometimes perform poorly, not only in terms of the technical quality of care [19–21], but also regarding key features of good primary care, such as patient-centeredness, accessibility [21,22], continuity of care [13,23], comprehensiveness, coordination, and community engagement [17,19]. For example, in Benin (West Africa), a study of physicians practising at primary care facilities showed that many PCPs struggle to provide high-quality primary care [24]. Moreover, the bulk of PCPs in most African countries work in urban areas and the private sector, which threatens equitable access to the services they provide [7,12]. These performance issues are alarming as they may lead to inefficiencies, given that PCPs remain a relatively rare and expensive source of care.

Fortunately, the current evidence indicates that, under certain conditions, the potential of PCPs can be unleashed to improve primary care in African countries. Indeed, the PCPs’ practices are diverse across various settings, and the literature reports positive outcomes for the practice models that are relatively better organised and guided by a set of clear principles and governance arrangements. This is, for instance, the case for family physicians who receive 2 to 4 years of postgraduate training in family medicine[16,17] or the "*Médecins Généralistes Communautaires”* who are community-based physicians serving in rural areas of some francophone African countries [7] or even physicians without specific postgraduate training but who have received some degree of governance guidance [25]. Therefore, understanding which factors influence PCP performance in a given context could help to guide better and improve their practices. This is especially important in countries like Benin, where primary care delivery largely relies on non-physicians and where PCPs still remain relatively scarce.

A review of the PCPs’ practices in sub-Saharan Africa, taking a health system lens [7], identified factors that may positively or negatively impact the performance of these practices. One key element is the PCPs’ professional identity - comprising values, motivation for primary care, and educational and professional trajectories - which is likely to shape their approach to primary care. Governance factors, such as policy guidance, role clarity, resource allocation, incentives, and accountability mechanisms, can also shape the performance of PCP practices. Additionally, the organisation of PCPs’ activities and their relationships with other primary care team members, local health authorities, community members, and key stakeholders are critical. Work environment factors (e.g., tools, equipment, infrastructure) and broader contextual elements (e.g., rural or urban setting, poverty levels, and overall health system performance) can further influence PCPs’ effectiveness and the performance of their practices. However, it remains essential to verify if these factors indeed impact the performance of PCP practices within each country’s specific context.

The literature examining the performance of PCP practices outside Africa also suggests similar factors, but it usually regroups these factors into physicians’ personal and professional characteristics on the one hand and organisational and systemic factors on the other [26]. The physicians’ characteristics most often pertain to their training and professional experience and, to a lesser extent, to their age and gender. The organisational and systemic factors include the organisation of the physicians’ practices (role and task distribution with other staff, for instance), their workload, adequate infrastructure and equipment availability, and the features of the local health system in which the PCPs’ practices are embedded. Organisational and systemic factors are reported to play a greater role in physicians’ performance (and the performance of their practices) than personal characteristics [26]. Yet, the specific factors (or combination of factors) that influence PCPs’ practices in sub-Saharan African countries are barely known [7].

This study aims to address this gap by exploring the factors that support the performance of PCPs’ practices in Benin. We define PCPs’ practices as the way the work of PCPs is organised, including their professional characteristics, activities, work settings, governance arrangements shaping their work, and interactions with other stakeholders. Performance is defined as the ability of PCP practices to deliver care that aligns with the core characteristics of quality primary care and to contribute to the effective functioning of their local health system. The performance of PCP practices thus includes both the individual performance of PCPs and their performance in collaboration with the primary care team.

The following questions guide our research: (1) What is the performance of selected PCPs’ practices in Benin? (2) Which factors are perceived by a diverse group of stakeholders to impact this performance? (3) Based on an analysis of empirical data, which factors appear most likely to support PCPs’ practices in providing quality primary care in Benin?

## Methods

### Study design

This is a multiple-case study using a combination of quantitative and qualitative methods. In this study, we defined the case as the performance of a given PCP’s practice.

### Selection of the study sites

To address the research questions, we needed to contrast various performance levels (good, average and poor) and observe multiple potential influencing factors. Therefore, we purposively selected 8 PCPs’ practices representing the study sites, using the PCPs as entry point. Each study site corresponds to one case. These sites were selected using a database from a previous empirical study [24]. This purposive sampling followed the steps described below.

#### Step 1: Defining sets of variables to use as criteria to predict the performance of PCPs’ practices

Because our objective was to include a range of cases representing different performance levels, we needed criteria to help us predict the performance of the practices of the PCPs in our database. This database comprises data from 150 PCPs [24]. A factor analysis of this data helped us to identify three sets of variables (see supplementary file 1). The first set includes variables related to the practice of quality improvement and public health activities. The second set includes variables related to the availability of essential health services (i.e., those defined in the Benin national guidelines as high-impact interventions) and the cost of PCPs’ services. The third set includes the PCP’s self-rated satisfaction. We used each set of variables as a criterion to predict the performance the PCPs’ practices. It is important to emphasise that we did not aim to construct a scientifically validated tool to assess performance but rather to find a pragmatic approach to establish a sufficient level of contrast between the cases included in this study.

#### Step 2: Predicting the performance of the PCPs’ practices based on each criterion

For each PCP, we calculated a score for each criterion, integrating the values of the variables composing this criterion. A score was considered low when below the mean score for a given criterion minus the standard deviation (score < μ- σ). A medium score was a score situated between the mean minus the standard deviation and the mean plus the standard deviation (μ- σ < score < μ+ σ). A high score was higher than the mean plus standard deviation (score > μ+ σ).

#### Step 3: Purposive selection of the PCPs’ practices to be included in the study

We purposively selected 8 PCPs whose practices would be included in the study. We aimed for a large mix: 2 PCPs with high scores for at least two criteria, 3 PCPs with medium scores across the criteria, and 3 PCPs with low scores for at least two criteria. In order to have a sufficiently large variation of cases, we also considered the gender and postgraduate training, if any, of PCPs, the institutional ownership of the health facility harbouring the PCPs’ practices, and the geographic position and the urbanisation level of the facility’s location.

Table 1 presents the characteristics of each of the 8 study sites who represent the 8 cases studied.

**Table 1:**
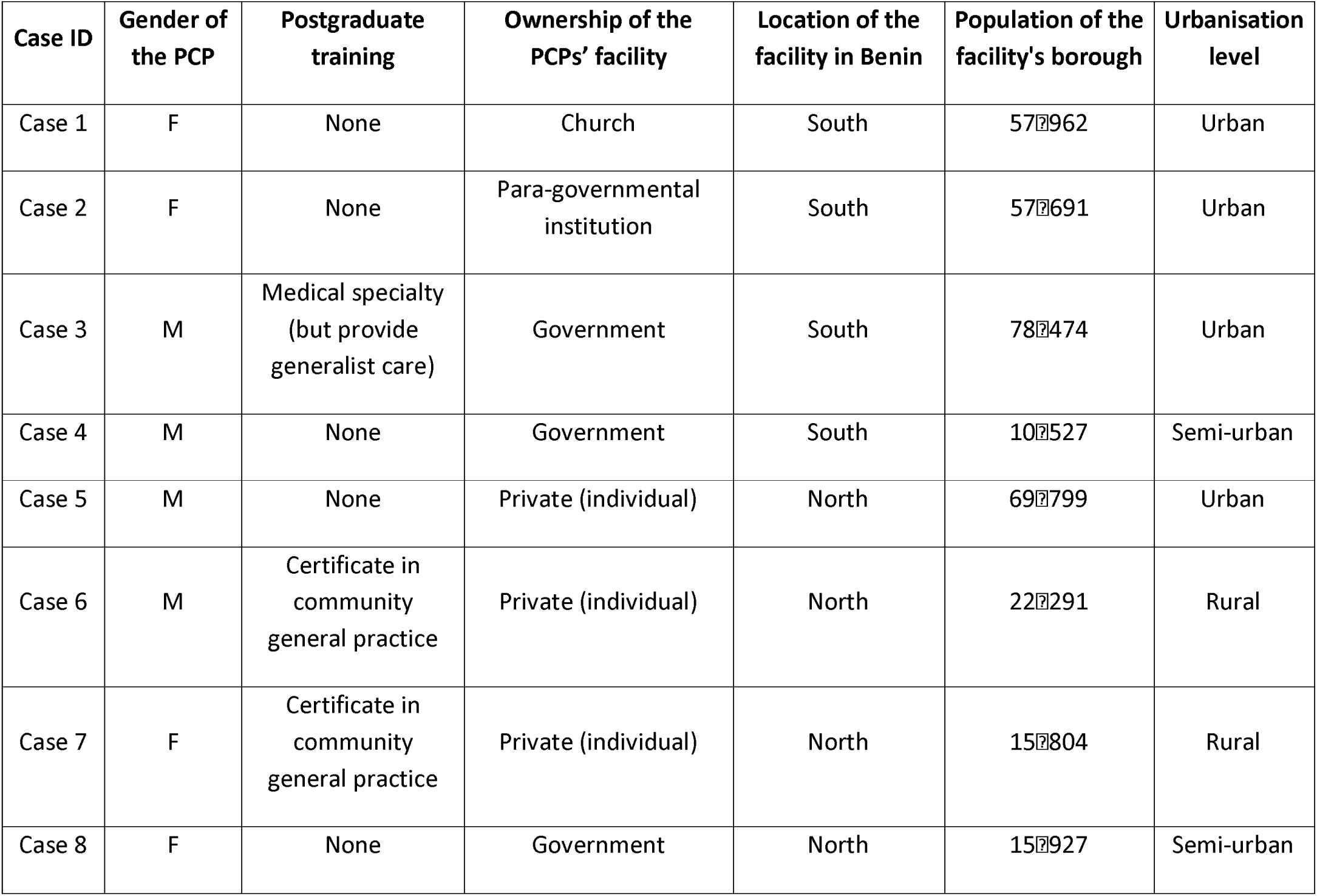
Characteristics of the study sites.

### Study settings

The study sites are spread across five health districts in Benin (figure 1). Three districts are urban: Cotonou 2-3 (Cot 2-3), Cotonou 5 (Cot 5) and Parakou-N’Dali (PN); and the two others are predominantly rural: Ouidah-Kpomassè-Tori (OKT), and Nikki-Kalalé-Pèrèrè (NKP).

**Figure 1:**
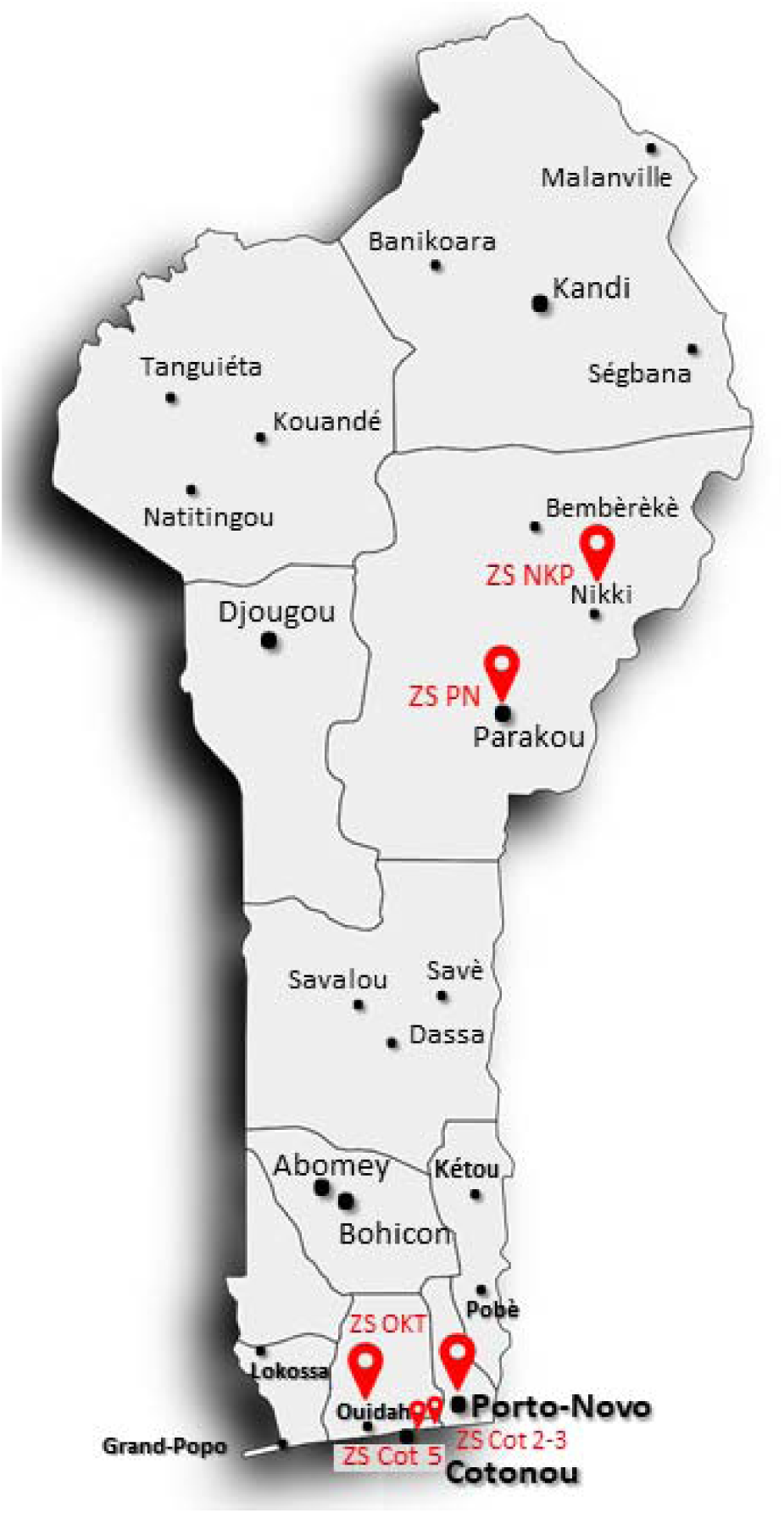
Benin’s health districts where the study was conducted.

### Conceptual framework

The conceptual framework (figure 2) used for this study was iteratively constructed based on the Donabedian model for evaluating the quality of medical care [27], the relevant literature on primary health care and health systems [28,29], and a scoping review previously conducted by the research team [7]. We also integrated into the framework the factors described in the literature (see above) as influencing PCPs’ practices [26].

**Figure 2:**
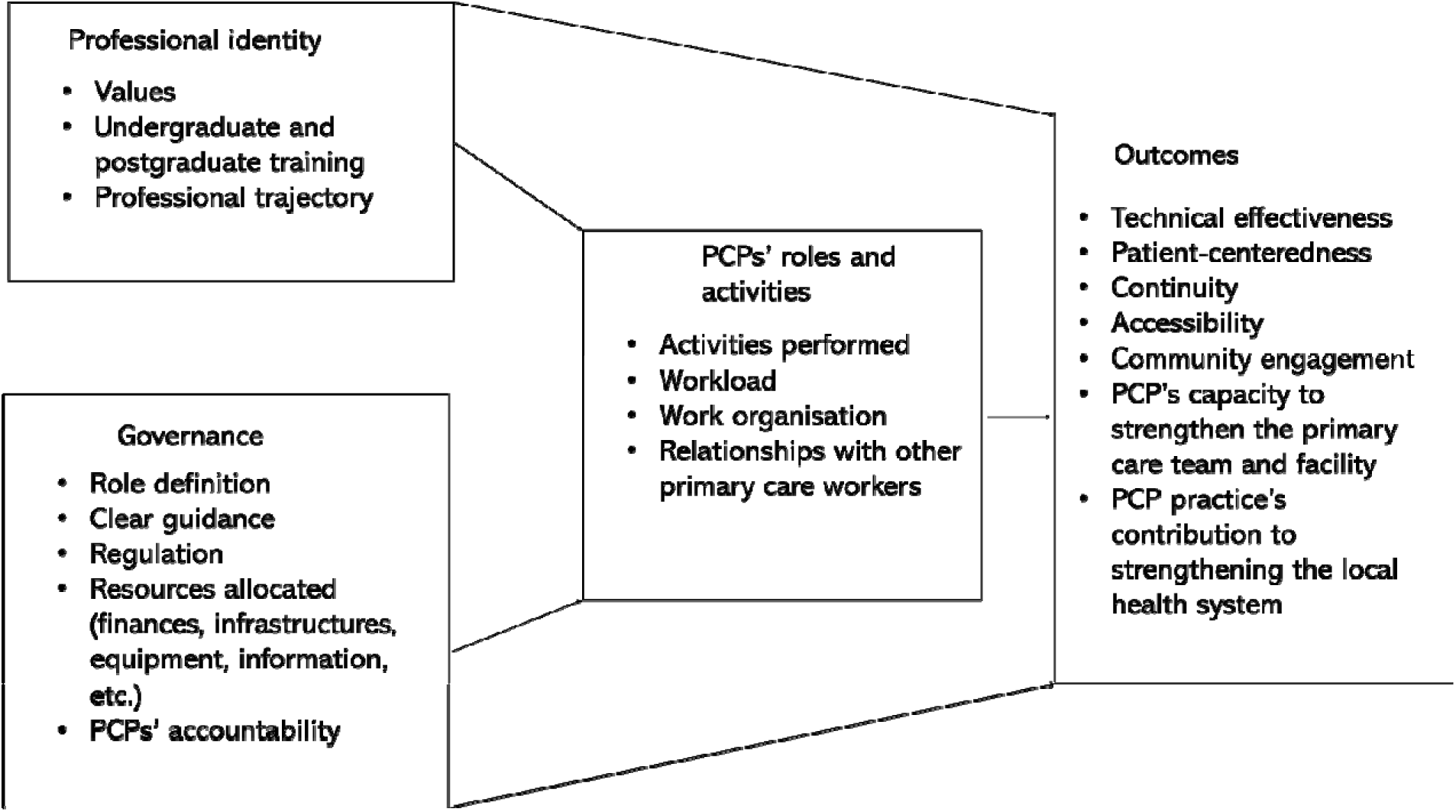
Conceptual framework.

In this framework, the outcomes box regroups the range of performance dimensions that we assessed in this study:

- technical effectiveness: which is the extent to which the care provided aligns with recognised biomedical standards of care.
- patient-centeredness: providing holistic care by considering not only the biomedical problem they bring in the consultation but also the psychological and social difficulties they may also have. It is also about dealing with the unique needs and preferences of the patients, and sharing information, power and responsibility with them.
- continuity: ensuring that the patients are followed over time by a clinician or a team of health care professionals and that there is effective and timely communication of health information.
- accessibility: the ease with which the population can initiate contact with the health professionals and use the services they need without any barrier such as geography, administrative hurdles, financing, cultural norms, and language.
- community engagement: working closely with the community to ensure the services provided are responsive to their need and collaborating with them to improve the health of the population.
- PCP’s capacity to strengthen the primary care team and facility: the extent to which the PCP’s work supports the performance of the primary care team and the quality of the services offered in the health facilities.
- PCP practice’s contribution to strengthening the local health system: the extent to which the PCPs’ practices contribute to the district health indicators and the overall performance of that district.

The first five dimensions are widely used to assess primary care performance [30–32], and we have chosen them because the ultimate goal of this work is to improve the contribution of PCPs’ practices to quality primary care in Benin and other African countries. Beninese stakeholders also perceived these dimensions as key to judging the performance of the PCPs’ practices. The PCP’s capacity to strengthen the primary care team and facility and the PCP practice’s contribution to strengthening the local health system are less commonly used to assess the practices of the primary physicians or primary care workforce in general. However, they were deemed important because the PCPs’ practices are meant to support the performance of the local health system and contribute to improving the health and well-being of the population. It is also important to note that all these performance dimensions, apart from the PCPs’ capacity to strengthen the primary care team and facility, include not only the individual performance of the PCPs but also that of the primary care team. Indeed, as PCPs rarely work alone in Benin, assessing their individual performance in isolation would not be appropriate.

From the literature [7,26,28,29], we identified a number of factors that can potentially influence the performance of PCPs’ practices. We regrouped them into three categories in the conceptual framework: first, elements pertaining to the professional identity of the PCPs; second, governance features of their practice; and third, the nature of activities conducted, and roles played in the local health system (see figure 2).

In this multiple case study, our goal was to test this conceptual framework based on the empirical data and retain the factors that actually influence the performance of the PCPs’ practices in the specific context of Benin.

### Data collection

Data collection took place from April to June 2022.

#### Quantitative data collection

The quantitative data addressed research question 1 and were collected through direct observations of curative outpatient consultations conducted by each of the eight PCPs whose practices were observed. A physician present in the consultation room performed the observations using a structured observation grid. This grid was developed based on two tools: the Global Consultation Rating Scale, a validated scale for assessing physician-patient communication and patient-centeredness [33], and a tool for evaluating the quality of clinical examinations following care protocols used in Benin [34]. These tools allowed us to quantitatively assess two key primary care dimensions: patient-centeredness and technical effectiveness.

We pre-tested the grid with five PCPs working outside our study area to assess its relevance and improved it based on their feedback. Additionally, a biostatistician reviewed the grid for further refinement. The physician observer was trained for this specific task, conducting ten joint observations with the principal investigator (KB) before starting the data collection. Their scores for each of the ten consultations were compared, and any discrepancies were discussed to ensure a consistent understanding of the grid.

Each PCP was observed over three consecutive days, chosen to capture the busiest periods as much as possible. Our goal was to observe at least 20 consultations per PCP, which was achieved for all except for case 7. The number of consultations observed for each PCP is displayed in Table 2.

**Table 2:**
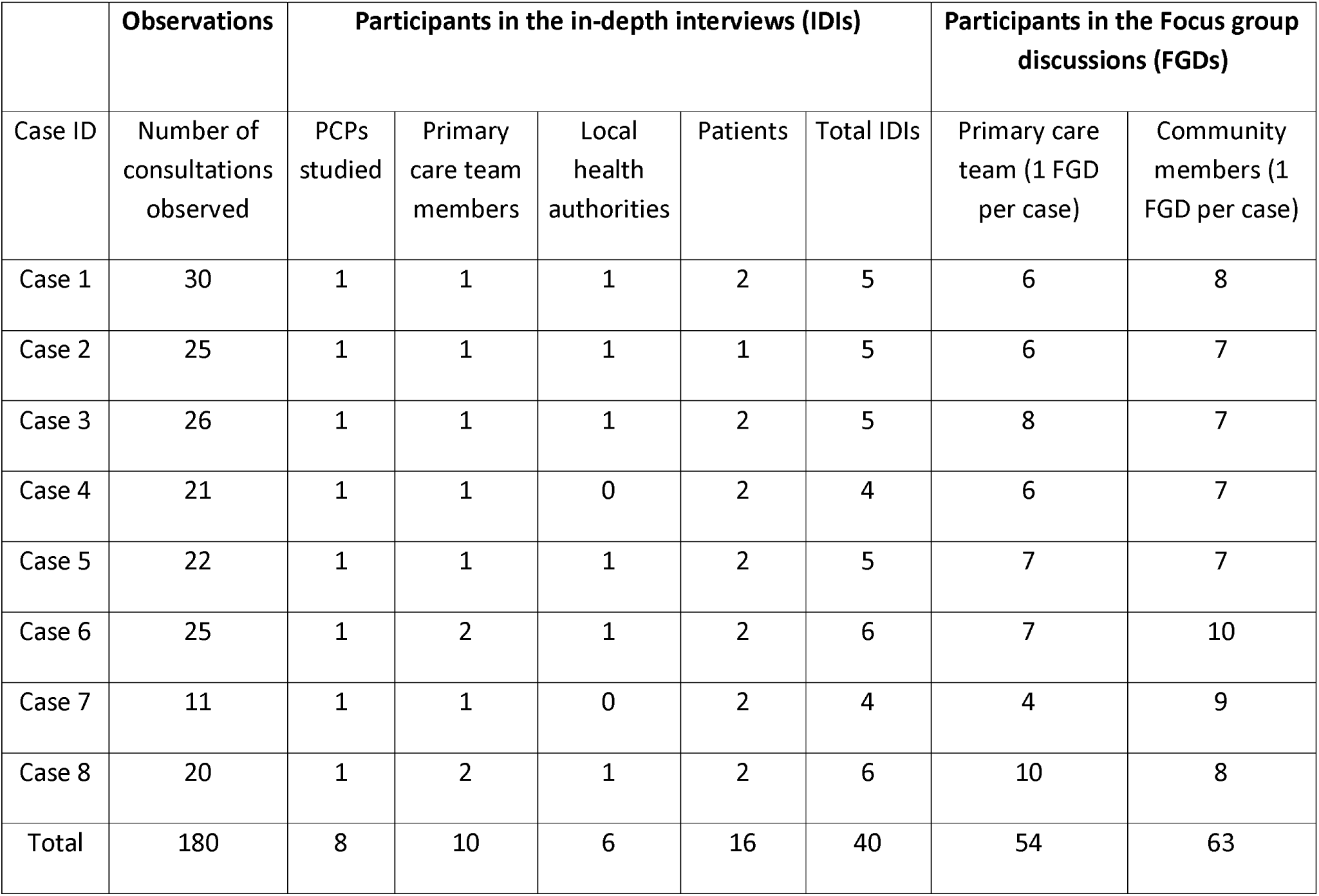
Number of consultations observed and number of participants in the interviews.

|  | Observations | Participants in the in-depth interviews (IDIs) |  |  |  |  | Participants in the Focus group discussions (FGDs) |  |
| --- | --- | --- | --- | --- | --- | --- | --- | --- |
| Case ID | Number of consultations observed | PCPs studied | Primary care team members | Local health authorities | Patients | Total IDIs | Primary care team (1 FGD per case) | Community members (1 FGD per case) |
| Case 1 | 30 | 1 | 1 | 1 | 2 | 5 | 6 | 8 |
| Case 2 | 25 | 1 | 1 | 1 | 1 | 5 | 6 | 7 |
| Case 3 | 26 | 1 | 1 | 1 | 2 | 5 | 8 | 7 |
| Case 4 | 21 | 1 | 1 | 0 | 2 | 4 | 6 | 7 |
| Case 5 | 22 | 1 | 1 | 1 | 2 | 5 | 7 | 7 |
| Case 6 | 25 | 1 | 2 | 1 | 2 | 6 | 7 | 10 |
| Case 7 | 11 | 1 | 1 | 0 | 2 | 4 | 4 | 9 |
| Case 8 | 20 | 1 | 2 | 1 | 2 | 6 | 10 | 8 |
| Total | 180 | 8 | 10 | 6 | 16 | 40 | 54 | 63 |

#### Qualitative data collection

The qualitative data were collected through individual in-depth interviews (IDIs) and focus group discussions (FGDs), utilising tailored interview guides that addressed all research questions. The interviews were conducted by the principal investigator (KB, a female physician) and two senior health sociologists (one male and one female).

For research question 1, we solicited respondents’ opinions on the performance of the PCPs and their respective teams. For research question 2, we explored respondents’ perceptions of the factors influencing the performance of the PCPs and their teams, both positively and negatively. For research question 3, the interview focused on eliciting themes which could possibly influence the performance of the PCPs’ practices, including their professional trajectories, relationships with team members, local health authorities, and community members, as well as their activities, the organisation of the primary care team, and the support they received. To enrich the findings for research question 3, we complemented the interviews with field observations, which provided insights into the PCPs’ work environment, available equipment and infrastructure, and the organisational aspects of their work.

For each case, key informants were purposively selected from various groups:

- All eight PCPs (IDI)
- Other primary care workers at the health facility, including physicians, nurses (mainly), midwives, nurse aides, and, where available, non-clinical staff (IDI and FGD). These interviewees were selected among the staff working closely with the PCP.
- Local District Health Authorities (IDI)
- Patients and community members, including local and religious leaders, community health workers, and representatives of community-based organisations (IDI and FGD). Patients were selected among those who attended the health facility during the period the research team visited the facility. For each PCP we selected at least one patient the PCP have consulted and where applicable one patient who was consulted by another primary care worker. The community members were selected with the help of a community health worker or a local leader among those who were well acquainted with the health facility where the PCP worked.

We adhered to the principle of maximum variation to ensure a diverse sample. All key informants provided information relevant to each of the three research questions. In total, we conducted 40 IDIs and 16 FGDs, comprising eight FGDs with 54 primary care workers and eight FGDs with 63 community members (see Table 2). The interviews with health workers were conducted at their health facilities, at a time convenient for them and in a private space. Interviews with community members were conducted within the community, also in private settings, most often in the participant’s home or, for FGDs, in a hut located in a community gathering space.

To gain a comprehensive understanding of PCP practices and performance, KB and the research assistants spent two months studying the PCPs’ practices in their specific contexts. Additionally, KB maintained contact with the PCPs for several months following data collection to address any follow-up questions as needed.

### Data analysis

#### Research question 1

We assessed the performance of each PCP practice, including both individual PCP performance and the performance of the primary care team.

Quantitative data were analysed using descriptive statistics to evaluate individual PCP performance. For each PCP, the mean communication score was calculated based on the Global Consultation Rating Scale [33], and we assessed the percentage of recommended clinical actions performed according to Benin’s care protocols.

Qualitative data were analysed for team performance. Most IDIs and FGDs were recorded and transcribed verbatim. For participants who declined the recording, we created a detailed summary immediately after the interview. Additionally, the research team maintained a daily log in Microsoft Forms, documenting all research activities and recording observation notes. Data analysis began during the fieldwork. The research team met daily after data collection to share their findings and discuss the performance of each case, based on the interviews they conducted and their observations. Divergences in opinion were noted, and additional data were collected the following days to help reach a consensus. After completing data collection for each case, one team member produced a memo summarising the team’s observations, which was then reviewed by the other members. All transcripts were uploaded to Dedoose [35] for thematic content analysis [36]. For coding, KB and the research assistants assigned interview excerpts to the corresponding performance dimensions. Following this, KB produced a summary for each case, by integrating the quantitative data and the respondents’ opinions. This summary was reviewed by the research assistants. The team then convened to assign a performance level (good, average, or poor) to each dimension for each case, taking into account all the data gathered and the analysis conducted (see supplementary file 2).

#### Research questions 2 and 3

We developed a coding tree based on factors indicated in our conceptual framework and field notes. Text fragments were assigned to the corresponding codes by KB and the research assistants during a data analysis workshop held in August 2022. Each fragment was then re-read to refine existing codes or identify additional or child codes. The coding tree was thus iteratively refined during the coding process with new codes progressively added (and other removed). Codes were then grouped into themes based on semantic similarities and co-occurrence patterns.

For research question 2, we summarised in a table the factors cited across the eight cases and stakeholder groups as significantly influencing the performance of the PCPs and their team, providing examples of positive and negative factors reported by respondents.

To address research question 3, we performed a cross-case analysis, which enables comparison of commonalities and differences in events, activities, and processes across cases (34). For this analysis, a matrix (supplementary file 3) was used to display all cases alongside the main themes identified as potential performance factors. We then examined the key aspects of each theme in greater detail and assessed its relationship to performance across cases. We concluded that a theme is probably an influencing factor for a specific dimension of PCP performance based on two conditions. First, the theme had to be consistently observed in cases with good performance on that dimension and either absent or rarely found in cases with poor performance. Second, a plausible causal pathway needed to be established to logically explain how this factor impacted case performance. In this paper, we summarise the most significant potential performance factors. The reasoning behind our selection of performance factors from the cross-case analysis is detailed in Supplementary File 4.

The cross-case analysis was performed together by all co-authors during several online discussions and reviews of preliminary findings.

### Ethics

This study received the ethical approvals of the Institutional Review Board of the Institute of Tropical Medicine of Antwerp in Belgium (N° 1545/21) and the local ethics committee for biomedical research of the University of Parakou in Benin (N°0513/CLREB-UP/P/SP/R/SA). We obtained informed consent from all participants, and the data were treated with strict confidentiality.

### Public involvement

PCPs and other stakeholders (including policymakers from the Ministry of Health) were not directly involved in setting the research questions. However, the research team held multiple discussions with these stakeholders about findings from prior studies on PCP practices. These engagements helped identify research gaps and informed the design of the current study. Furthermore, the results of this research were presented to policymakers, NGOs, PCP associations, other health worker organizations, and patient associations in October 2022.

### Reflexivity statement

This research partnership prioritised equitable collaboration between Benin-based and European researchers. The study was locally led by KB, a Beninese public health researcher, who drove all stages from design to dissemination. Senior researchers (DMZ in Benin; JDL/BC in Belgium) provided mentorship.

Capacity strengthening included: training provided by the first author (KB) to Beninese research assistants in qualitative methods and the use of tools like the Dedoose software, and supporting KB’s PhD development, including (but not limited to) coaching for academic writing and advanced data analysis methods. Funding also supported KB’s work environment (e.g., computer, office supplies) and her affiliated Beninese research centre.

Data analysis and interpretation were collaborative, with regular meetings (starting during data collection) to ensure inclusion of local perspectives. The multidisciplinary nature of the research team (primary care researchers, public health researchers, clinicians, and sociologists) enriched the interpretation of findings through diverse viewpoints. Findings were shared with Benin stakeholders in October 2022 and were instrumental in the development of a policy framework to guide PCP practices in Benin.

While the authorship includes only one woman (KB), her leadership role advances gender equity in African health research. The study received Benin ethical approval, and data were managed with strict confidentiality to protect participants and local researchers.

## Results

### Performance of the eight PCPs’ practices

Figure 3 displays the mean communication scores and the mean percentage of recommended clinical actions performed for each of the PCPs (based on quantitative data collected through consultation observations).

**Figure 3:**
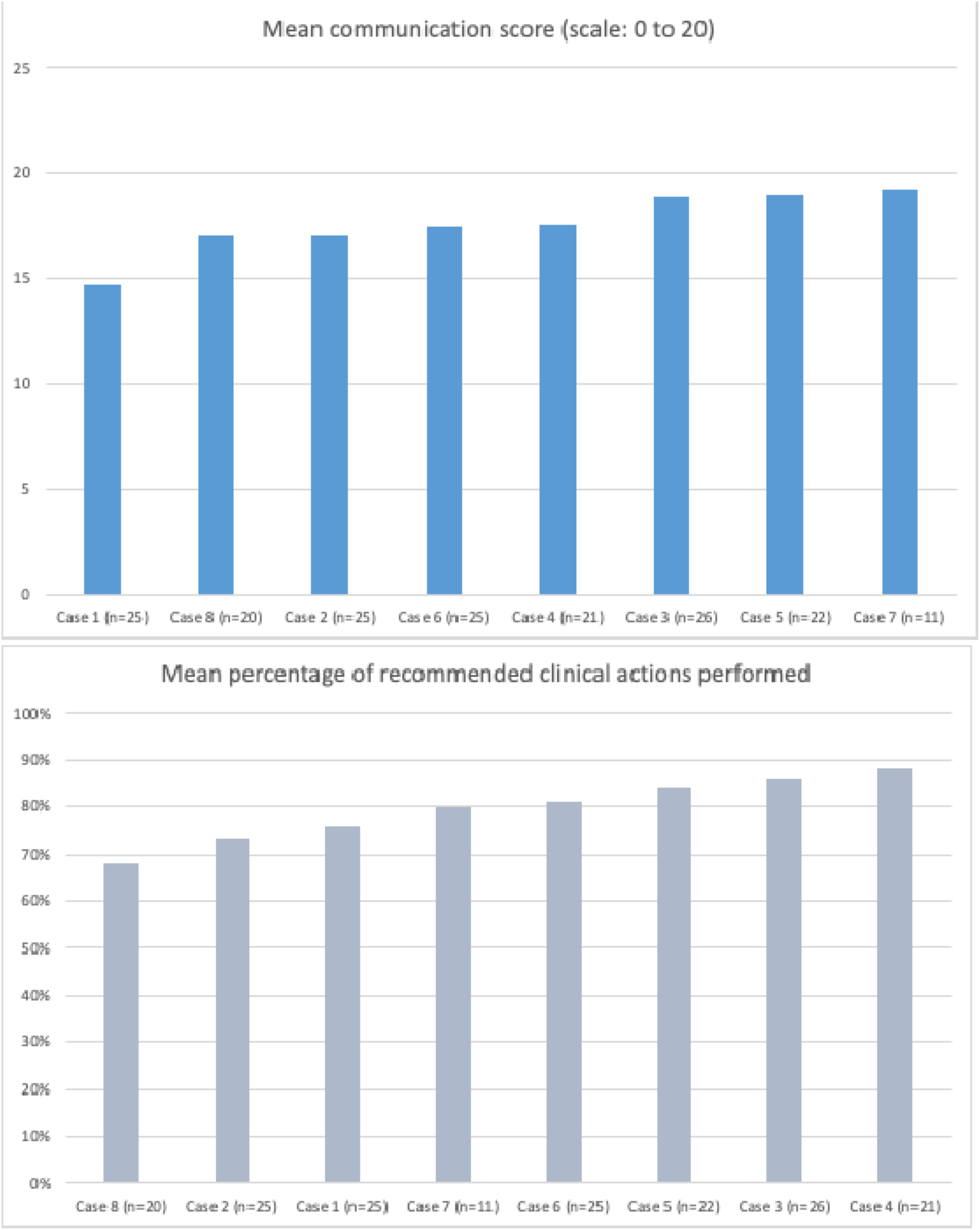
Mean quality scores of the consultations observed for the PCP in each case.

Table 3 below summarises the performance of the 8 cases, as assessed by the study team by integrating the quantitative and the qualitative results and triangulating the views of various stakeholders. The detailed analysis that led to this summary is provided in Supplementary File 2.

**Table 3:**
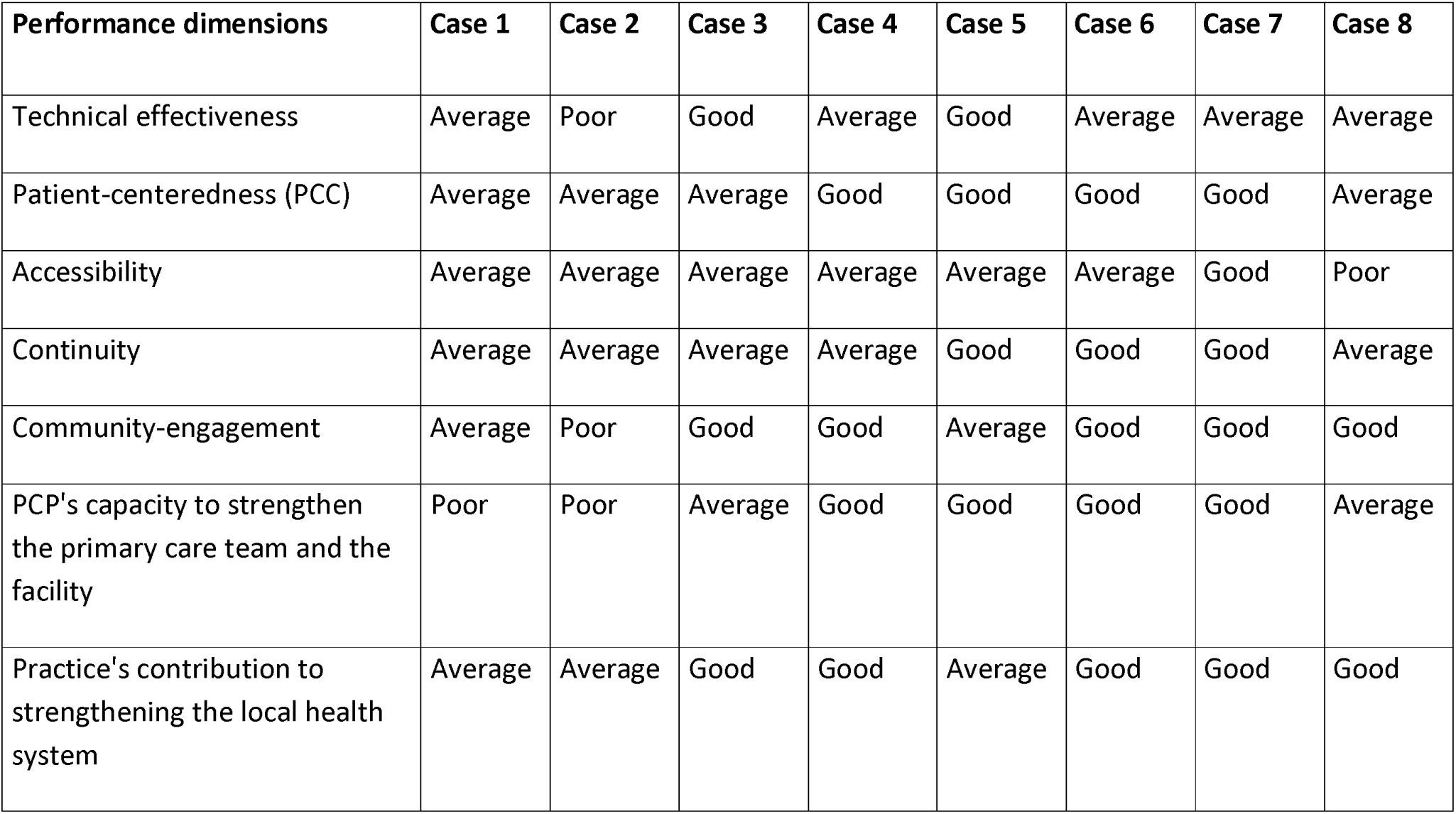
Performance of PCPs’ practices, based on quantitative and qualitative findings integration.

The results above show that cases 1 and 2 represented the weakest performance for all dimensions assessed. The PCPs in these cases had the lowest communication scores (14.69 and 17.08, respectively) and the lowest percentages of recommended clinical actions (68% and 73%, respectively). Similarly, based on stakeholders’ perceptions and observations, the performance in these two cases was systematically poor or average throughout all dimensions.

The other six cases were more difficult to categorise. When we look at the individual performance for technical effectiveness and patient-centred care, assessed through the consultations’ observations, case 8 showed low scores. However, the qualitative data indicated good performance of this PCP’s practice in terms of community engagement and contribution to strengthening the local health system. Cases 3 and 5 appear to be the best-performing cases when considering individual performance during observations. Indeed, even if they did not have the best score for any of the two indicators, they both had high scores for both (see Figure 3). The qualitative data also confirmed this finding. However, when we integrate the other dimensions and the performance of the primary care team as a whole (see supplementary file 2), the performance in cases 3 and 5 is rather average. For instance, patient centeredness of care provided in case 3 is average because of poor welcoming and communication with patients by the primary care team (except the PCP). Moreover, the accessibility is limited because the PCP is regularly absent. As for case 5, its accessibility is limited because of the high care costs. There is also a lack of genuine community engagement beyond top-down designed sensitisation campaigns. The contribution of the case 5 to strengthen the local health system is difficult to appreciate because of the absence of a clearly defined population of responsibility and limited coordination with other health system actors.

Case 6 and 7 however appeared to be the strongest performance. Case 6 showed an average performance for both quantitative indicators (17.47 for the communication scores and 81% for the percentage of recommended clinical actions). The best-performing case for communication was case 7 (score of 19.28), but it was only average for the other indicator (80%). Similarly, the best-performing case in terms of complying with recommended clinical actions was case 4 (88%), but it was only average for the other quantitative indicator (score of 17,63). However, the qualitative data revealed that in cases 6 and 7, the performance was better in more dimensions than in the other cases.

### Factors perceived by various stakeholders as key influencers of the performance of PCPs’ practices

The factors that the stakeholders most frequently mentioned were related (in descending frequency order) to (i) equipment, infrastructure, and utilities; (ii) support of the primary care team to the PCP; (iii) PCPs’ intrinsic motivation and values; (iv) PCPs’ knowledge and competencies; (v) fulfilment of the PCP’ personal needs; (vi) support from health authorities, peers, local government authorities and/or community members; (vii) workload that PCPs face; (viii) general context elements of the locality where the PCPs’ practice is situated; and finally (ix) financial resources available at the PCPs’ health facilities.

Table 4 provides illustrations of how these factors influence the performance of PCPs’ practices, as reported by the stakeholders.

**Table 4:**
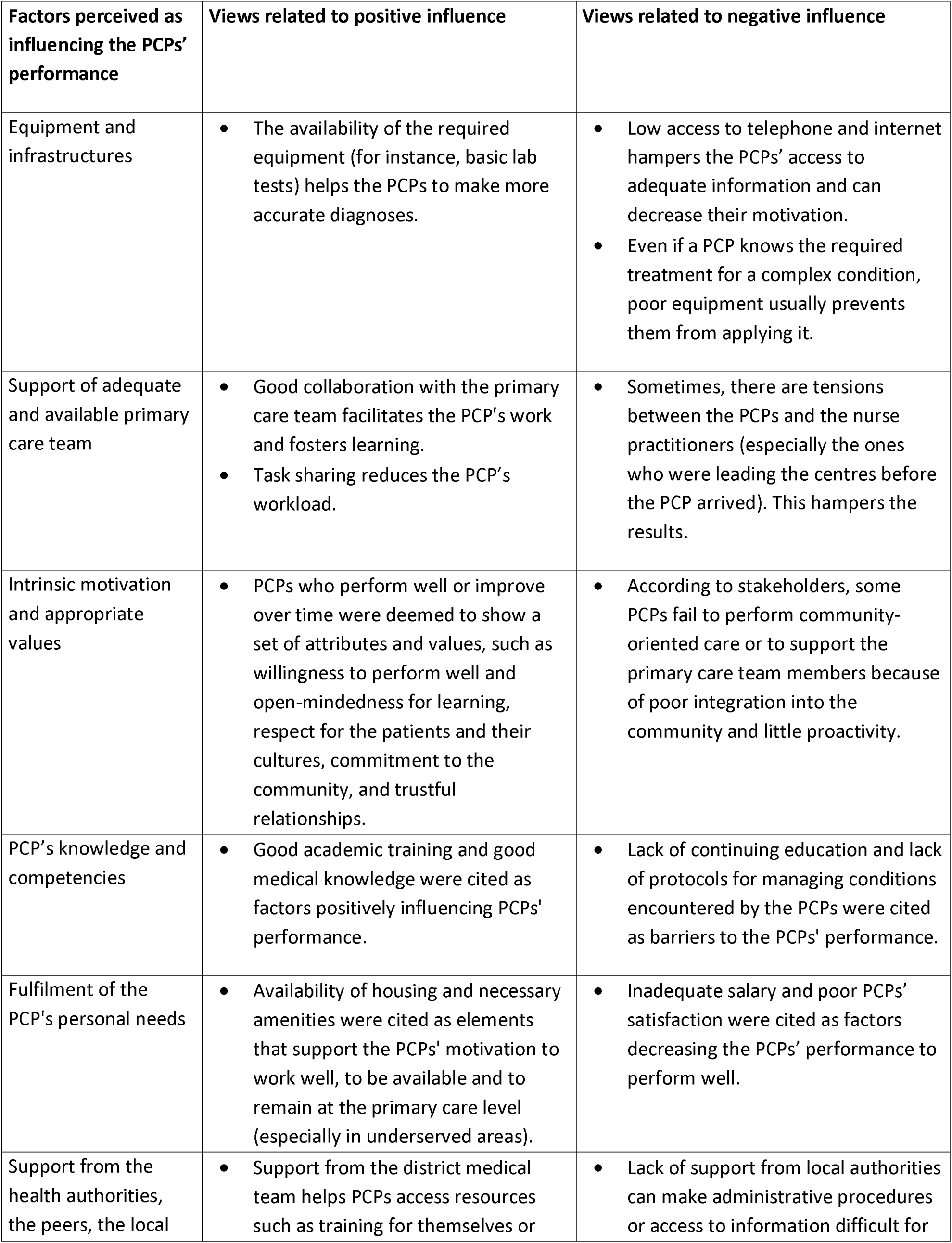
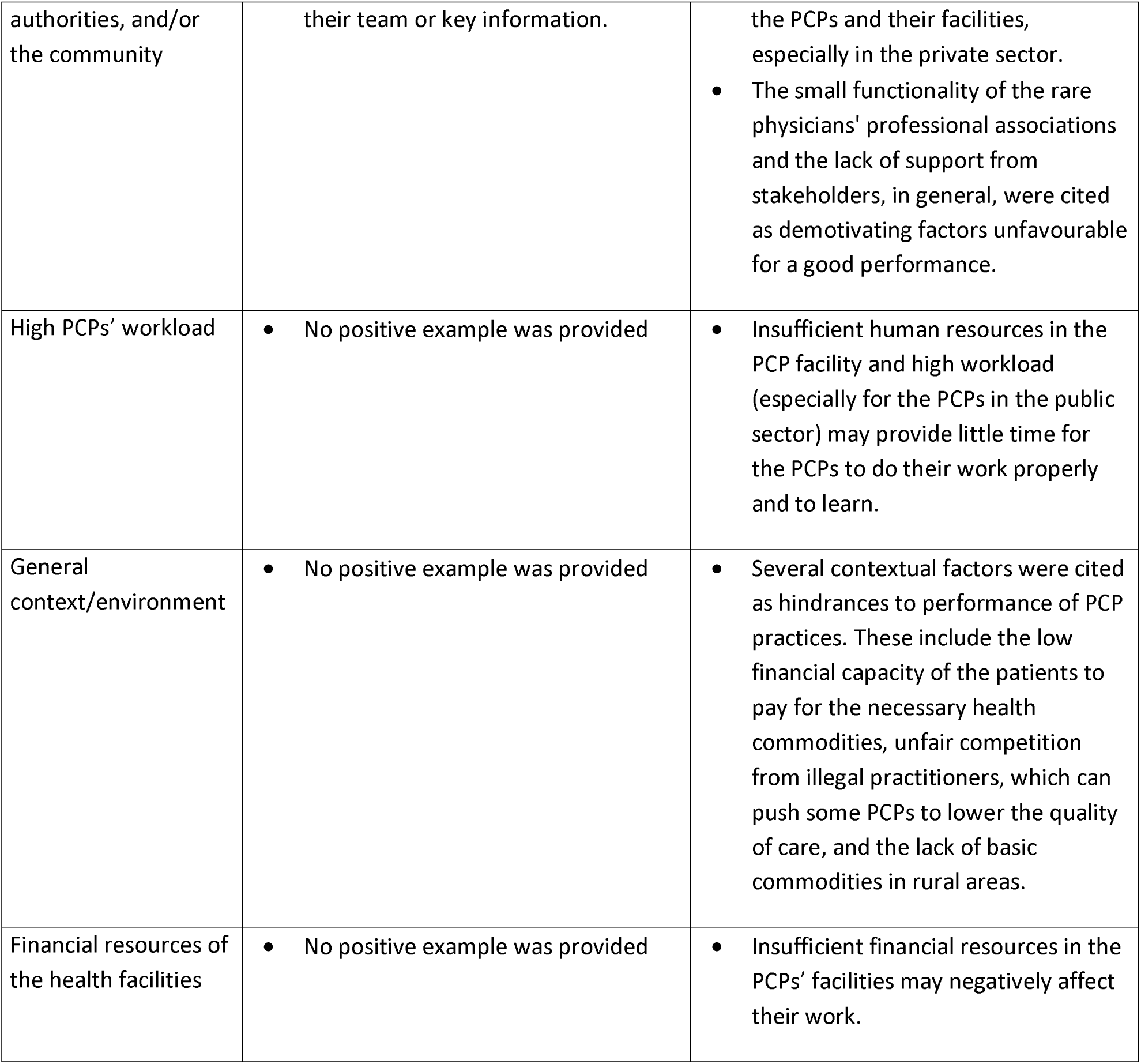
Factors the stakeholders perceive as influencing the PCPs’ performance.

### Factors highlighted by the cross-case analysis as likely to contribute to PCPs and their team providing quality primary care in Benin

Based on stakeholders’ perceptions, field observations, and initial data analysis, we identified a set of factors that could potentially influence the performance of PCP practices. A cross-case analysis of the eight cases explored these factors and concluded that the following are likely to influence the performance of PCPs’ practices: (i) the values underpinning the PCPs’ practices; (ii) the PCPs’ preparation to practice at the level of the first line; (iii) the support provided to the PCPs by the hierarchy, peers or professional associations; (iv) the leadership mandate given to the PCPs and the degree of autonomy allocated to exert it; (v) the modalities of financing of the PCPs’ practices; (vi) the accountability mechanisms in place to support PCPs’ practices; (vii) the extent of collaboration with community leaders and other key stakeholders; (viii) the PCPs’ relationship with the rest of the primary care team and their leadership style; and (ix) the context in which the PCPs’ practices are embedded, especially whether this practice is in urban and rural area, and the ownership of the practice by the State or not. The detailed analysis of all potential factors across the 8 cases leading to retain the above-mentioned factors is available in the supplementary files 2 and 3. However, we provide below an illustration of the reasoning leading to these findings.

The cases 1 and 2 represented the weakest performance in our sample. In these cases, the PCPs had little preparation to practice at the first line level, the mandate and roles assigned to them were limited to patients’ consultations, and they had limited autonomy for influencing anything in their health facilities. They did not have any incentive or requirement to go into the community, and they worked in silos or in minimum collaboration rather than in teams with the other primary care team members (especially case 2). On the contrary, in cases 6 and 7, which performed better than others in several dimensions, the PCPs were very well prepared through short postgraduate training and field coaching. As the owners of their health facilities, they had a leadership mandate and the necessary autonomy to exert this mandate. They also signed a charter with their professional association, the health authorities, and the local authorities through which they pledged to serve a well-defined population and work together with the local stakeholders. In these cases 6 and 7, the PCPs lead the primary care teams, but they exert a person-centred leadership with good collaboration with the other primary care team members and particular attention to the needs of these team members. Regarding the values, we found, for all cases, a commitment to serve the community and a degree of professionalism. However, differences were found across the cases for other values, which appears to impact performance. For example, in the practices where altruism and commitment to help people were more developed (cases 1, 6 and 7), the financial accessibility was better. On the contrary, in sites where these values were lacking (case 5) or only officially stated (cases 2, 3,4, and 8), the financial accessibility was either average or bad. Similar patterns were found for the support provided to the PCPs, the modalities of financing the PCPs’ practices, the accountability mechanisms, and the context.

Even though equipment and infrastructure are the most often mentioned factors by all types of stakeholders, we could not find enough evidence to conclude that these factors influence the performance of the practices studied. Neither could we conclude that continuing education and fulfilment of the PCP’s personal needs would influence the performance of the PCP. We did not analyse the impact of the overall health regulation on the PCPs’ practices because this regulation is the same from one case to another, and we couldn’t find differences among the cases.

Table 5 summarises our key findings.

**Table 5:** Summary of the key findings from the cross-case analysis.

| Corresponding aspect of the conceptual framework | Observed factors | Summary of the analysis |
| --- | --- | --- |
| Professional identity | Values | • The values that guide the PCPs' practices influence the outcomes of these practices.<br>• The nature of the values determines which performance dimension is influenced and whether this influence is positive or negative. For example:<br>- Altruism and commitment to helping people positively influence the financial accessibility of the services provided.<br>- Hierarchical culture negatively impacts the PCPs' capacity to strengthen the primary care team.<br>• Some proclaimed values (for instance, the public service orientation in the public health facilities) were good, but they are not supported by the incentives and job descriptions provided to the PCPs. |
| Governance | Preparation of the PCP to practice at the first-line | • PCPs who were prepared (i.e. empowered in terms of knowledge and skills, psychological preparation to work in resource-constrained environments, and preparation for the specificities of the primary care practices) to fulfil their roles at the first line tend to perform better in the area in which they were capacitated.<br>• From the examples in our study, this preparation can occur during undergraduate training in some medical schools, during postgraduate training, or on the job through coaching at the start of employment.<br>• The aspects in which the PCPs are capacitated determine the dimensions in which they will be performant. For example, one of our cases was mainly performant in technical quality and facility management; these were the dimensions in which he was very well prepared. |
| Governance | Continuing education | • There is a general lack of continuing education among the cases studied.<br>• Our findings do not support a relation between continuing education and the performance of the PCPs' practices. |
| Governance | Equipment and infrastructure | • There is generally poor equipment and infrastructure in the practices studied, but our findings do not support equipment as a factor determining the PCP's performance. |
|  |  | • Our findings do not support the fact that equipment and infrastructure impact the performance of the PCPs' practices. |
| Governance | Support from the hierarchy, peers or associations | • Having support positively influences the performance of the PCPs who benefit from it. This support helped the PCPs to have a better understanding of the roles expected of them, to have access to some necessary resources, to get feedback on their performance, and to have psychological resilience in difficult situations. This, in turn, improved their performance.<br>• PCPs who received support in our sample received it either from the hierarchy, their peers, or professional associations.<br>• The performance dimension influenced by the support provided to the PCPs depends on the type of support provided (thus, the support should be tailored to what is expected from the PCPs). |
| Governance | The leadership mandate given to the PCP and the degree of autonomy allocated to exert it. | • The PCPs who have the mandate to lead their health centre or primary care team took action to strengthen this team and to improve their health facility.<br>• However, our findings suggested that the leadership mandate has a greater chance to support the PCPs' performance when it is associated with a certain degree of autonomy. |
| Governance | Modalities of the financing of the PCPs' practices | • The reliance on the income generated by the health services reduces the financial accessibility in some cases because the PCPs' practices needed to increase their costs to ensure their financial viability. This also led to over-prescription in other cases, thus reducing the quality of care.<br>• The existence of financial support from a third party (NGO or church, in cases 1, 6 and 7 for instance) helps to maintain the costs low, thus improving financial accessibility.<br>• Some strategies may help PCPs' practices mitigate their financial burden: the use of generic medicines, targeting a high number of consultations instead of a high price for one consultation, reducing the open hours, and a good accounting system supported by a professional accountant.<br>• The financing modalities can improve the PCPs' accountability and performance. For example, relying on the income generated by the health services may improve the technical quality of care and patient-centeredness if the patients have the possibility to choose other health practitioners (as in case 5). |
| Governance | Fulfilment of the PCP's personal needs | • The fulfilment of the PCP's personal needs appeared to increase the motivation and retention in the workplace for some PCPs. However, others could achieve good performance in various dimensions even with only an average satisfaction related to their personal needs. |
|  |  | We did not have enough evidence to conclude that it improves the performance of PCP practices. |
| Governance | Regulation of the PCPs' practices | In Benin, there are rules and laws that exist to regulate the practices of physicians and health workers in general (medical code of ethics, decrees regulating the private practice of health professionals, law for the people's health protection in Benin, etc.).<br>We did not analyse this factor because the regulation does not differ from one case to another. |
| Governance | Accountability mechanisms | Effective accountability mechanisms (including a clear definition of the PCPs' roles, the clear definition of the deliverables or indicators expected from them and an effective way to assess these deliverables or indicators) positively influence the performance of PCPs' practices.<br>The dimensions of performance influenced by the accountability mechanisms depend on the expectations set for the PCPs and the possibility of properly assessing (or measuring) the attainment of these expectations.<br>Patients' and community members' capacity to hold the primary care teams accountable can positively influence the performance of the PCPs' practices. This capacity comes either from their financial capability (such as in case 5) or from the strength of the community leaders (cases 6 and 7 for instance). |
| Roles and activities | Collaboration with community leaders and other public officials | PCPs' practices where we found an active collaboration with community leaders and other key stakeholders were more likely to have better community engagement and a better contribution to strengthening the local health system.<br>Collaboration with community leaders and other key stakeholders can also help the PCPs' practices to have access to some resources and feedback on their performance. |
| Roles and activities | Relationship with the rest of the primary care team and leadership style of the PCPs | A strong collaboration between the PCPs and the rest of the primary care team is associated with a person-centred leadership style from the PCPs, which positively influences the performance of the PCPs' practice, especially for the PCPs' capacity to strengthen the primary care team.<br>This positive influence is achieved through better teamwork, knowledge sharing, and the development of shared values.<br>A hierarchical relationship and a command-and-control style of leadership, and a relationship where the PCP and the rest of the primary care team members work in silos appear to not support a good performance.<br>A hierarchical relationship and a command-and-control style of leadership may support good performance of PCPs' practice, but only if the PCP has control over the hiring of primary care team members. This was observed for instance in cases 3 and 5, where both PCPs exhibited this leadership style, and both had good individual performance. However, in case 5, the primary care team members performed better than in case 3, likely because the PCP in case 5 had greater control over hiring these team members. |
|  |  | • Allowing nurse practitioners to provide patient care contributes to increasing the accessibility of care for the population. This is achieved in two ways: the nurses can compensate for the absence of the physicians, and having nurses that consult reduces the number of physicians necessary, thus the cost for the facilities and the patients. |
| Roles and activities | Workload | • Difficult to appreciate a possible impact of the number of patients on the PCPs' performance, as the average number of patients seen daily was relatively low, ranging from 4 to 10.<br>• Even in the case of limited availability due to non-clinical tasks (especially for the PCPs in the public sector), this was compensated by the nurse practitioners. Therefore, it was also difficult to appreciate the impact of multiple non-clinical activities on the performance of the PCPs' practices. |
| Not directly related | Context in which the PCPs' practices are embedded | • Several contextual aspects were found: insufficient financial capacities within the communities, urbanisation level, ownership of the facility, and recurrent health systems issues such as a weak referral system or medicine stockout at the national level.<br>• In our study, the contextual factors that influence the PCPs' performance were:<br>- Urbanisation level: Some conditions appear easier to find in rural areas than in urban areas, such as a good relationship with the community leaders or a family-like relationship between the PCP and the rest of the care team. Lab tests, branded medicines and private health insurance were more available for the cases located in urban areas. This increases the availability of services but also provides more incentives for over prescription, as seen in some cases. Moreover, the competition with other centres tends to be higher in urban areas, which led some of the cases to propose more and more diagnosis tests and sophisticated equipment to outdo other facilities.<br>- the ownership of the facility: In facilities owned by the State, the degree of autonomy of the PCPs and ability to make some decisions is lower than in the facilities owned by the PCPs themselves. |

## Discussion

### Main results and comparison with international literature

#### Assessing the performance of the PCPs practices

In this study, we used seven dimensions to evaluate the performance of the PCPs’ practices: technical effectiveness, patient-centeredness, continuity, accessibility, community engagement, PCP’s capacity to strengthen the primary care team and facility, and PCP practice’s contribution to strengthening the local health system. This holistic view of performance makes the evaluation complex, and simpler approaches could be advocated for routine monitoring. However, our holistic approach enabled us to get an in-depth understanding of the performance of PCPs’ practices and to identify issues that might not be uncovered with the analysis of only one dimension. For instance, in our study, the two cases with the best technical effectiveness were average for other performance dimensions, which impeded the overall quality of the care received by the patients. Measuring the performance of the health workers’ practices in primary care systems should thus include various dimensions, reflecting the final outcomes expected from these systems. This multidimensional assessment is a strong recommendation of the PHC measurement framework published by WHO in 2020 [31] and other works [37,38]. Unfortunately, many studies on health workers’ performance assessed one or very few dimensions, usually the technical effectiveness for specific health conditions [39–41].

Another important fact is that the PCPs’ performance can hardly be disentangled from that of their team and the health facility in general. This is particularly true in contexts like Benin and many other African countries where the PCPs usually work together or along with nurse practitioners. This is why we analysed the performance of the PCPs’ practices as a whole, instead on focusing on the individual performance of the PCPs. The case study approach we used also allowed us to analyse the performance of the PCPs’ practices within their context and understand how the PCP’s performance relates to the performance of the rest of the healthcare team and to the patient’s experiences. For example, we found that some PCPs might have excellent individual performance, but the overall performance of their practices (which includes their own performance and the one of their team) was rather average. In other cases, the individual average performance of the PCPs was compensated by a good performance of the primary care team, resulting in good patient’ experience and good contribution to strengthening the primary care team. Our approach to studying the performance of the PCPs’ practices aligns with the recommendations in the literature, according to which one should not only focus on measuring and improving the performance of individual health workers but also consider the performance of the health care team [26,37,41].

#### Factors influencing the performance of PCP practices

Nine factors emerged from this multiple-cases study as likely to influence the performance of PCPs’ practices in Benin:

- One factor related to the professional identity of the PCPs is the values supporting the PCPs’ practices.
- Five are related to the governance of the PCPs’ practices: the PCPs’ preparation to practice at first- line, the support provided to the PCPs from the hierarchy, peers or professional associations, the leadership mandate given to the PCPs and the degree of autonomy allocated to exert it, the modalities of the financing of the PCPs’ practices, and the accountability mechanisms in place to support the PCPs’ practices.
- Two are related to the roles and activities of the PCPs: the PCPs’ relationship with the rest of the primary care team and their leadership style and the PCPs’ collaboration with community leaders and other public officials.
- One transversal factor: the context in which the PCP’s practices are embedded.

For the values, both the analysis of the stakeholders’ perceptions and the cross-case analysis showed that they relate to various performance dimensions. Professional values are indeed cornerstones of the identity of any profession [42], and they are reported to be important for health workers’ performance [42]. However, as clearly illustrated in some of the cases included in our study (table 5) and other studies [43], professional values should not be taken for granted, and it is not enough to articulate them in official documents. It is important to ensure that the PCPs and other primary care workers adopt the values that would support the performance expected from them. The few existing postgraduate training programs for PCPs in sub-Saharan Africa [44,45] and some undergraduate programs [46] include socialisation of the trainees for the core PHC and family medicine values, such as social justice, people-centeredness, community orientation, etc. However, our study shows that values can also be shared through interactions with peers within professional associations or coaching from supervisors. This provides opportunities for PCPs already working and countries without a postgraduate training program.

As for the PCPs’ preparation, providing the PCPs with the necessary knowledge and skills to practice in primary care settings was among the top five factors cited by the stakeholders we interviewed. Scholars also pinpointed physicians’ knowledge and skills as key factors in determining their performance [37,46]. However, knowledge and skills should help the PCPs achieve the core primary care functions [31] and be geared towards the people’s needs at the primary care level (as opposed to a hospital setting) [47]. Other aspects of the PCPs’ preparation include preparing them psychologically to work in resource-constrained environments and improving their soft skills (leadership and communication skills, for example) to embrace the various roles expected from them, to work within a team and to deal with the community leaders [8,37,48]. Our study showed that the PCPs who got such preparation tend to perform better. Similar to the values, this preparation can be provided through various channels, including undergraduate training, postgraduate training, or coaching by professional associations or supervisors.

Another factor highlighted by the stakeholders’ interviews and the cross-case analysis is the support received by the PCPs. Studies in both hospital and primary care settings found that support from the health facility managers helps to implement best practices [41], motivates the health workers for good performance [37,38], and facilitates access to necessary resources to provide care of good quality [41]. Peer support is also reported in the literature as a powerful means to enhance health workers’ performance [49].

Regarding the leadership mandate, the PCPs in our sample who were given this mandate make visible efforts to improve the quality of care and the availability of resources in their health facilities. We also found that among PCPs with a leadership mandate, those with the necessary autonomy were more able to make the necessary decisions and hold the primary care team accountable for good performance. Leadership is a key function in the health system [31,50]. In a primary care team, this function can be fulfilled by any cadre [48]. However, many stakeholders in Africa view the PCP as the natural leader of the primary care team [8,48,51], and our study shows that this is also the case in Benin. Furthermore, to strategically position PCPs to optimise their contribution to primary care performance, it may be an effective strategy to formally assign them the leadership mandate, along with the necessary skills to execute this role effectively.

Concerning the financing modalities of the PCPs’ practices, the cross-case analysis showed that a lack of financial support is likely to lead to increased costs for the patients and poor quality. Providing financial support to the PCPs’ practices can thus contribute to improving their performance (and their financial sustainability) [52]. For instance, we found in this study, as in other work [37,38], that the opportunity to receive funding (or risk of losing it) was an incentive to be more responsive to the patients. However, this financial support (and the PCPs’ payment mechanisms) must be carefully designed and geared towards the results expected from the PCPs [2,37]. It should avoid the perverse effects sometimes related to financial incentives by integrating a mix of provider payment mechanisms [2] and being combined with other types of incentives.

The accountability mechanisms in place to support the PCPs’ practices strongly appeared in the cross-case analysis as influencing the PCPs’ practices. Effective accountability mechanisms include a clear definition of the PCPs’ roles (this was, for example, the case for the PCPs’ practices in the public sector in our sample), a clear definition of the expected results (including the population for which they are accountable), effective ways to assess and share these results and clear consequences when these results are achieved or not [53]. The literature also reports the need for the PCPs and the primary care teams in general to be accountable [37,38,48]. However, our study highlighted that accountability mechanisms can exist and be enforced but not support the right results. For instance, the PCPs in the public sector in our sample were held accountable for the income generated by their health facility, but this does not necessarily support the core functions of primary care. Therefore, the accountability mechanisms for PCP practices in Benin should be built around the end results expected from the primary care system.

The PCPs’ relationship with the rest of the primary care team (mentioned by the stakeholders as the support of an effective and available primary care team) is another performance factor found by this study. Like in other research [2,31,41,54], we found that the overall performance of the PCPs’ practice is likely to be better if the PCPs work in cohesion with other primary care professionals (especially nurse practitioners). When such cohesion exists (rather than working in silos or within hierarchical relationships), there are better chances for the primary care team to have shared values, share knowledge and work together towards a shared objective [41]. Moreover, in our study, we found that when there is teamwork, the non-physicians are able to provide good quality care to the patients, with the support of the PCP, even when the latter is not present. This teamwork requires a person-centred leadership style from the PCP, one that fosters shared objectives, promotes mutual understanding, motivates team members, and builds team spirit [38,48].

A good collaboration with community leaders and other public officials also appeared in the cross-case analysis and stakeholders’ interviews to facilitate the performance of the PCPs’ practices. Indeed, when such good collaboration exists, the community leaders usually help the PCPs understand the needs of the communities they serve and build the necessary trust for the use of the health services by these communities. We also found some cases where the community leaders helped the PCPs access financial or material resources or provided feedback on their practices, thus contributing to the overall performance of these practices. Engaging with local stakeholders to understand key issues and design and implement solutions for these issues has long been proven to be an essential factor for improving performance and ensuring the success of health interventions [31,38]. However, this factor is more commonly cited when referring to health system governance or facility management [31,38]. Our study shows that engaging key stakeholders in the community is also important for the delivery of good quality primary care services and the performance of the primary care teams.

The last factor emphasised by both the cross-case analysis and the stakeholders’ interviews is the context in which the PCPs’ practices are embedded. We found that this context influences the relative importance of other factors on PCPs’ performance. For instance, a PCP with a person-centred leadership style appears to be even more important in the public sector than in the private sector. Indeed, the PCPs in the public sector don’t always have the necessary decision space to enforce orders given in a command-and-control way. Also, lab tests, branded medicines and private health insurance were more available for the cases located in urban areas. This increases the availability of services, but it also provides more incentives for over prescription. The socio-cultural and economic context indeed influence the practice of any profession, and this should be considered when designing interventions to improve health workers’ performance [26,37,38]. The level of urbanisation warrants particular attention as the share of the urban population is quickly growing worldwide. In Benin, the urban population constituted 49.5% of the country’s population in 2022, and this proportion is projected to be 65.45% by 2050. In Western Africa, 63.80% of the population is expected to live in urban areas by 2050 [55]. However, some positive factors seem easier to get in rural areas (for instance, a good relationship with community leaders). Therefore, planning for primary care services and PCPs’ practices should carefully consider how to avoid losing the advantages linked to rural areas and avoid or work around the shortcomings inherent to urban settings.

#### Factors with insufficient evidence regarding a potential influence on the performance of PCPs’ practices

Even though they were cited by the stakeholders as influencing factors, we did not find sufficient evidence in the cross-case analysis to conclude that the following factors influence the PCPs’ performance in Benin: the equipment and infrastructure available for the PCPs’ practices, the continuing education, the fulfilment of the PCPs’ personal needs and the workload of the PCPs.

The lack of equipment and infrastructure and the quasi-inexistence of a well-structured continuing education system are common situations found across Benin’s health system. In our study, we did indeed observe this lack of resources and continuing education in most of our cases, but we realised that those who performed well managed to overcome these issues. For example, in some cases, PCPs managed to ensure good continuity of care despite the lack of digital patient records, which could have facilitated their work. Also, almost all the cases that displayed good results with patient-centred care managed to do so despite physical spaces that were not always appropriate. Finally, PCPs who had a leadership mandate, the necessary will or creativity, the necessary autonomy, or the right relationships with community leaders managed to gain access (even if temporarily) to resources for their work. Similarly, some of the cases in our sample were able to compensate for the lack of continuing education, thanks to peer support. Therefore, despite the importance of adequate equipment [26] and continuous training [26,40], our results show that much can be done even with limited resources and other factors that can help to have a good performance.

Other studies have also reported successful experiences of primary care provision in resource-limited settings and even in situations of fragility [38,56]. This opens great possibilities in countries similar to Benin, with an urgent need to improve the quality of care for their population but where the socio-economic situation will probably not allow the PCPs to get the same resources as PCPs in better-resourced countries.

Regarding the workload, some studies found that a high number of patients negatively influences the physicians’ performance [26]. In our study, it was difficult to appreciate such influence, as the average number of patients seen daily by the PCPs was relatively low. Also, the PCPs with a high non-clinical workload in our sample were less available for patient care. This was compensated by nurse practitioners, and we did not find enough evidence to conclude that it influences the performance of the PCPs’ practices.

### Implications for practice

#### Contribution to international knowledge

The factors identified in this study align with existing literature, yet nuances emerged specific to the context of Benin and relevant to other low-resource settings. For instance, we found that several approaches can be used to prepare PCPs for working at the primary care level, including undergraduate and training postgraduate training, as well as coaching and on-site support from peers and supervisors. In countries lacking postgraduate for PCPs, undergraduate programs could be strengthened to better integrate essential primary care competencies, with increased clinical rotations in primary care facilities instead of predominantly hospital settings. Additionally, implementing onboarding programs at the start of a PCP’s career and establishing structured coaching systems—where senior PCPs and experienced health managers provide guidance—could enhance practical skill acquisition and support good performance.

Another important contribution of this study is the finding that while resource limitations (such as insufficient equipment, infrastructure, or continuing education) can hinder performance, they can be partly offset by strong leadership and support from peers, communities, or health authorities.

The study also highlighted structural factors, such as PCP preparation and context-specific shared values, which extend beyond individual physician characteristics yet hold significant potential to shape the performance of PCPs’ practices. These structural factors appear to be root causes of the underperformance seen in PCPs’ practices and, more broadly, in primary care systems. Identifying these factors provides essential insights for improving the contribution of PCPs’ practices to high-quality primary care in Benin.

Finally, this study provides empirical evidence that, when combined with existing research, can inform a theory on supporting the performance of PCPs’ practices in Benin, potentially forming the basis for a policy framework and guiding future efforts to analyse and improve the performance of PCP and other primary care workers in Benin and similar contexts. Although this study did not provide a sufficient foundation to build a full theory, it enabled us to develop a refined conceptual framework that can be further strengthened through additional research in varied settings.

#### A refined conceptual framework for PCP performance

As described in the methods and results sections, the cross-case analysis revealed causal pathways linking identified factors to the performance of the PCPs’ practices. These pathways indicate that the performance factors identified in this study are interlinked, with some acting as intermediary factors that mediate the influence of other factors on the performance of PCPs’ practices. Additionally, the pathways include key processes, such as resource mobilisation and PCPs’ activities, that play a role in shaping performance outcomes (see Figure 4).

**Figure 4:**
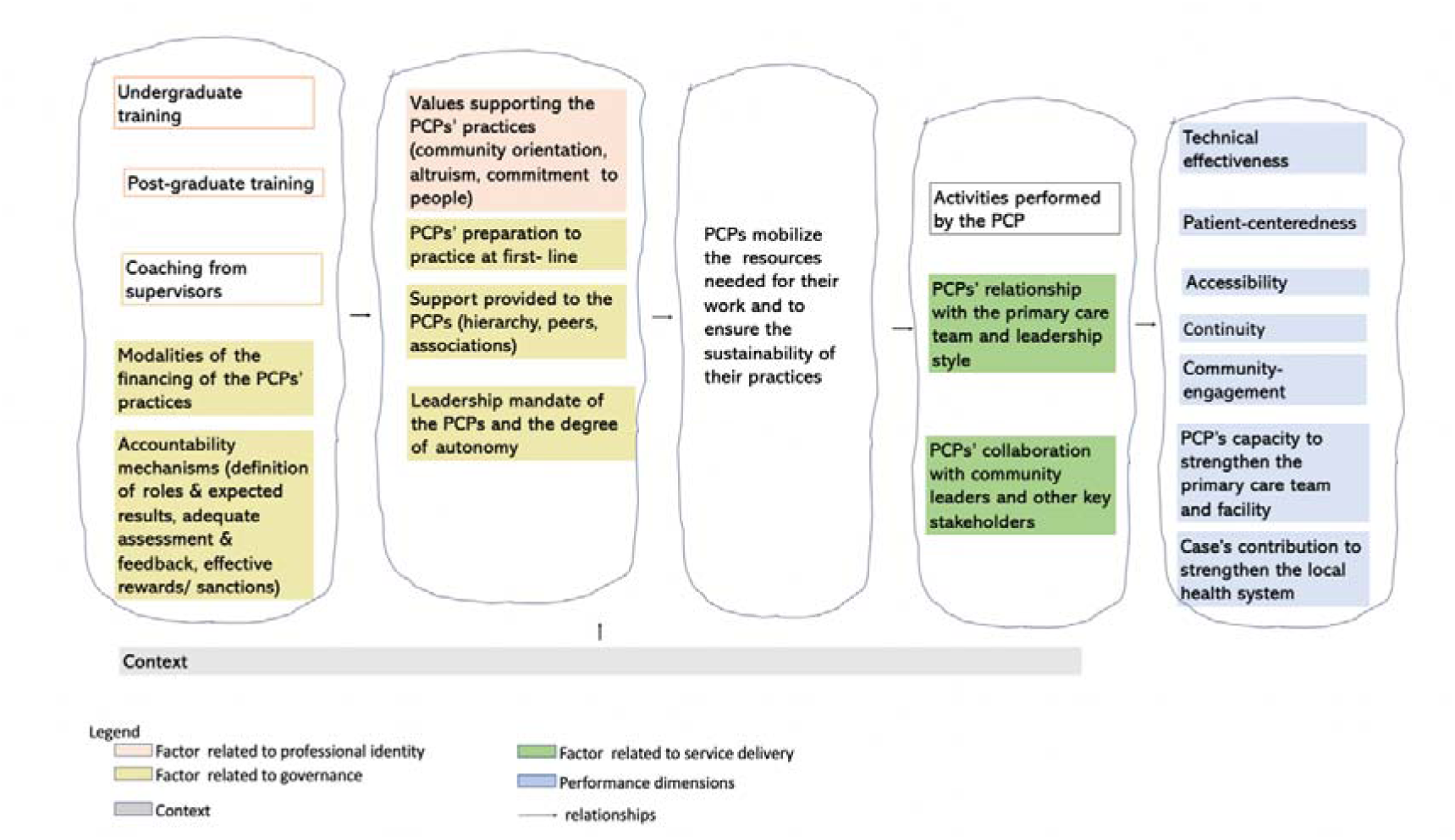
Pathways to PCPs’ performance.

The text below outlines the causal pathways illustrated in Figure 4.

" If we offer adequate undergraduate training and adequate postgraduate training and/or the right coaching, the right financing modalities, and effective accountability mechanisms, we will have PCPs with the right values and the right preparation, the necessary support, and a leadership mandate with the autonomy to exert it. These PCPs will then be equipped to mobilise the resources needed for their work and to ensure the sustainability of their practices. This, in turn, can enable PCPs to carry out their activities in good collaboration and teamwork with the rest of the primary care team members and engage with community leaders and other key stakeholders. All of this can ultimately lead to good performance of the PCPs’ practices, meaning that the PCPs and their teams provide care that is technically effective, patient-centred, accessible, and community-oriented. Additionally, PCPs could be able to strengthen their teams, and the PCPs’ practices could contribute effectively to reinforcing the local health system. These pathways to performance will also be shaped by the context".

It is also essential to note that the relationships between performance factors and the performance of the PCPs’ practices may be reciprocal. High-performing practices could enhance factors such as effective collaboration with community leaders, increased support from health authorities and communities, improved accountability, and stronger relationships within the primary care team.

Further studies are necessary to elucidate better the causal direction of the relationships between the performance of PCPs’ practices and the performance factors identified in this study.

Figure 5 presents the refined conceptual framework derived from these descriptions of causal pathways. In this refined framework, the conceptual framework introduced in the methods section was adapted to emphasise the performance factors identified through the cross-case analysis. Factors highlighted by key informants, which were not confirmed by the cross-case analysis, have also been retained as performance factors, but an interrogation mark has been placed beside them to indicate that they require confirmation. Additionally, elements of the initial conceptual framework that were not identified in this study have been removed. We have also started putting bidirectional arrows and put some factors before others to reflect the relationships found.

**Figure 5:**
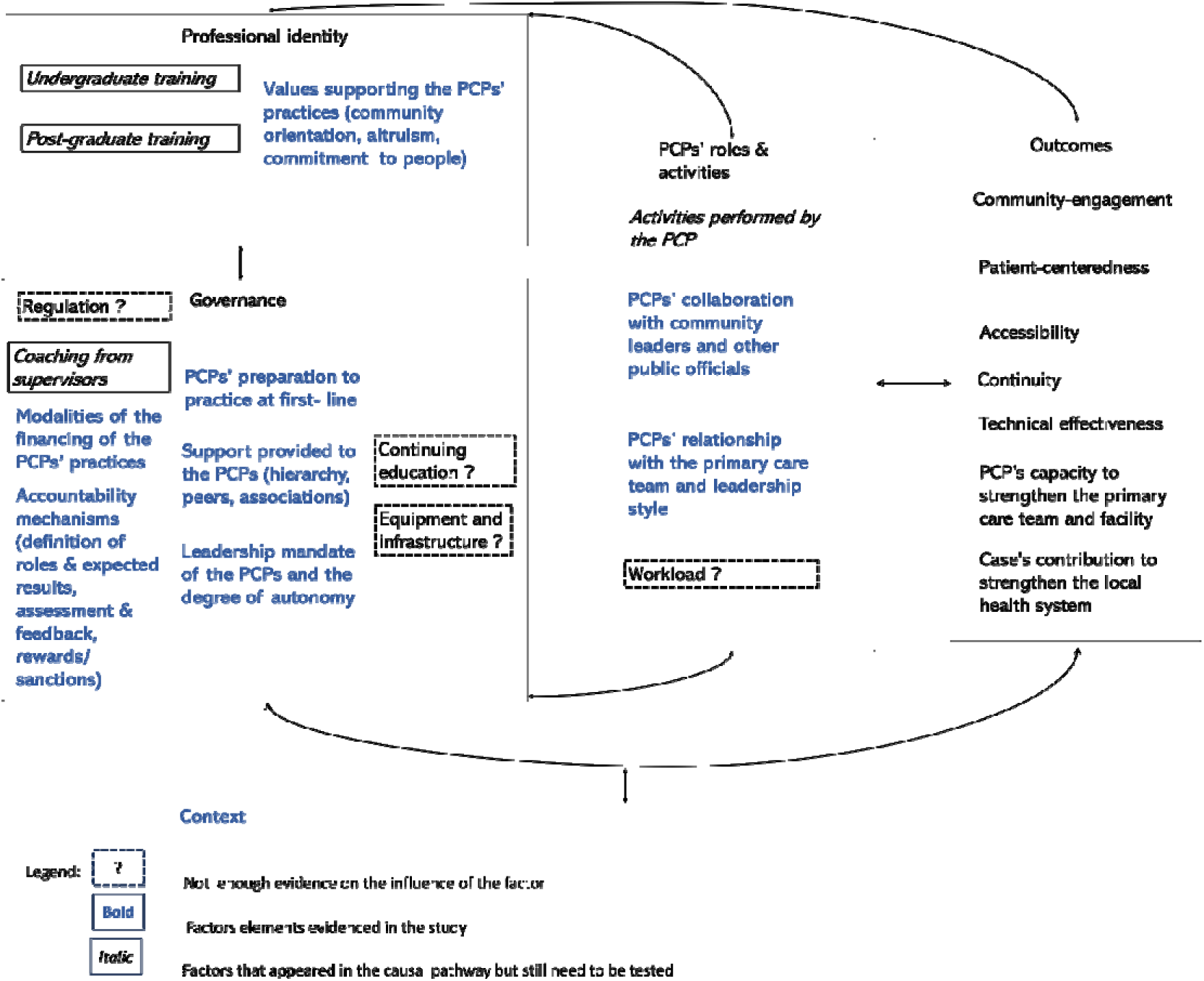
Refined conceptual framework for performance of the PCPs’ practices.

### Limitations and strengths

We used validated tools for assessing the individual performance of the PCPs but had only one observer to assess the consultations (instead of at least two as recommended for such assessment [33]). It was difficult to have more than one observer because of the small consultation rooms and some reluctance from the PCPs and patients. This may have reduced the reliability of our findings. However, the triangulation between quantitative and qualitative data helped us to mitigate these insufficiencies. We also discussed the assessments between the observer and principal instigator. Another limitation is the low number of cases included in the multiple case study. There is thus room to study the range of factors influencing PCPs’ performance on a larger sample and to test empirically the refined conceptual framework derived from our study. Furthermore, although using multiple dimensions allowed for a deep and nuanced understanding of PCP practice performance, it also made the cross-case analysis and the identification of performance factors more complex. This complexity may have led to misinterpretation of some links between the factors and performance, or even introduced some confirmation bias. We sought to limit these risks through multiple team discussions during the data analysis process. We also invited one experienced health system researcher to provide feedback on the first draft of this paper. This feedback was used to refine the paper.

Finally, the assessment of team-level performance was based on qualitative data rather than quantitative measures using standardised or validated tools. While we attempted to reduce subjectivity through data triangulation, qualitative observations, and structured team discussions to rate each case, this approach may still limit the robustness of our findings.

Future research could further explore the performance of PCP practices in Benin and similar settings, as well as the factors influencing this performance. Building on the performance dimensions and factors identified in this study, researchers could also consider using more computational approaches, such as Multi-Criteria Decision Analysis [57], to rank the performance of practices and the relative importance of influencing factors.

Despite these limitations, the study has several strengths. We achieved a reasonable level of variation in our sites in terms of performance, geographic localisation, urbanisation level, gender, and institutional ownership. This variation provided opportunities to observe similarities and differences across cases and note how a given factor would play a role (or not). Moreover, the principal investigator and research assistants spent two months interviewing and observing the primary care teams and other key stakeholders, which improved their understanding of the PCPs’ practices. Finally, we succeeded in conducting 56 interviews, which allowed us to reach saturation of data.

## Conclusion

By triangulating data from stakeholders’ perceptions, field observations of PCP practices, and evidence from the literature, we identified key factors shaping the performance of PCPs’ practices in Benin: (i) the values supporting the PCPs’ practices, (ii) the PCPs’ preparation to practice at first- line, (iii) the support provided to the PCPs from the hierarchy, peers or professional associations, (iv) the leadership mandate given to the PCPs and the degree of autonomy allocated to exert it, (v) the modalities of the financing of the PCPs’ practices, (vi) the accountability mechanisms in place to support the PCPs’ practices, (vii) the PCPs’ relationship with the rest of the primary care team and their leadership style, (viii) the PCPs’ collaboration with community leaders and other public officials, and (ix) the context in which the PCP’s practices are embedded.

Although the case-study methodology used in this study limits generalisation, the factors identified in this study can be tested in future research on PCP performance in other African countries. Furthermore, this study highlights the close link between factors influencing the performance of PCPs and the specific expectations and working conditions they face. The findings were instrumental in refining a conceptual framework for PCP performance, representing an initial step toward developing coherent policies on the organisation and regulation of PCP practices in Benin and other West African countries with similar health systems.

## Data Availability

All data produced in the present study are available upon reasonable request to the authors.

## List of abbreviations

Cot 2-3: Cotonou 2 and 3
GP: General practitioner
IQR: Interquartile ranges
MGC: Médecin généraliste communautaire
NKP: Nikki-Kalalé-Pèrèrè
OKT: Ouidah-Kpomassè-Tori
PN: Parakou-N’Dali
PCP: Primary care physician
PHC: Primary health care
qual (in lowercase letters): Non-dominant qualitative strand in a mixed method study
QUAN (in uppercase letters): Dominant quantitative strand in a mixed method study

## Declarations

### Ethics approval and consent to participate

This study was conducted under the ethical approval N° 1545/21 issued by the Institutional Review Board of the Institute of Tropical Medicine of Antwerp and the ethical approval N° 0513 of March 1 2022, issued by the local Ethics Committee for Biomedical Research of the University of Parakou (Benin). Written consent was obtained from the participants, and we managed the data with strict confidentiality.

### Consent for publication

Not applicable.

### Availability of data and materials

The data supporting the findings are included in the tables in this published article and its supplementary information files. More information is available from the corresponding author (KB) if needed and on reasonable request.

### Authors’ contributions

KB co-conceptualised the study, coordinated the data collection, curated the data, performed quantitative and qualitative data analysis, and produced the first draft of the manuscript. JDL and BC co-conceptualised the study, supervised the research process, contributed to the data analysis, and reviewed the manuscript drafts. DMZ supervised the research process (including protocol writing and data collection) and reviewed the manuscript drafts. All authors have read and approved the manuscript.

## Acknowledgements

We sincerely thank Dr Marinelle Houngbedji, Mrs Jeanne Madindé and Mr Maxime Mama for their valuable support in the data collection and data analysis. We also thank Prof Bruno Marchal for reviewing this manuscript and greatly enhancing its quality through his comments.

## Supplementary files

Supplementary file 1: Factor loading table for the three set of variables used to construct the criteria to approximatively appraise the PCPs performance

Supplementary file 2: Detailed performance analysis for each case

Supplementary file 3: Cross case analysis matrix

Supplementary file 4: Detailed narrative of the cross-case analysis

## References

1. United Nations General Assembly. Transforming our world: the 2030 Agenda for Sustainable Development. [Accessed on October 30, 2024]. United Nations: 2015. Available from: https://sdgs.un.org/2030agenda#:~:text=We%20are%20resolved%20to%20free,a%20sustainable%20and%20resilient%20path.

2. Expert Panel on Effective Ways of Investing in Health. Definition of a frame of reference in relation to primary care with a special emphasis on financing systems and referral systems. Brussels : European Commission; 2014. https://health.ec.europa.eu/system/files/2019-11/004_definitionprimarycare_en_0.pdf.

3. Shi L. The impact of primary care: a focused review. Scientifica (Cairo) 2012:432892. doi: 10.6064/2012/432892.

4. World Health Organization, United Nations Children’s Fund. Operational Framework for Primary Health Care: Transforming Vision Into Action. Geneva: World Health Organization; 2020.

5. World Health Organization. Global strategy on human resources for health: Workforce 2030. Geneva: WHO press; 2016.

6. Bosongo SI, Mukalenge FC, Tambwe AM, Criel B. Les médecins prestataires à la première ligne des soins dans la ville de Kisangani en République Démocratique du Congo1: vers une typologie. Afr J Prm Health Care Fam Med. 2021;13(1), a2617. 10.4102/phcfm.v13i1.2617

7. Bello K, De Lepeleire J, Kabinda MJ, Bosongo S, Dossou JP, Waweru E, et al. The expanding movement of primary care physicians operating at the first line of healthcare delivery systems in sub-Saharan Africa: A scoping review. PLoS One. 2021; 16(10):e0258955. doi: 10.1371/journal.pone.0258955

8. Eyal N, Cancedda C, Kyamanywa P, Hurst SA. Non-physician clinicians in sub-Saharan Africa and the evolving role of physicians. Int J Health Policy Manag. 2015;5(3):149–53. doi: 10.15171/ijhpm.2015.215

9. Willcox ML, Peersman W, Daou P, Diakité C, Bajunirwe F, Mubangizi V, et al. Human resources for primary health care in sub-Saharan Africa: progress or stagnation? Hum Resour Health. 2015;13(76):1–11. doi: 10.1186/s12960-015-0073-8.

10. Desplats D, Koné Y, Razakarison C. Pour une médecine générale communautaire en première ligne. Med Trop. 2004;64(6):539–44. French. doi: 10.1055/s-0029-1237558.

11. Ministère de la santé du Bénin. Politique nationale de santé 2018-2030. Cotonou: Ministère de la Santé du Bénin; 2018. French. https://files.aho.afro.who.int/afahobckpcontainer/production/files/VF_PNS__19_01_2020_VF.pdf

12. Bello K, De Lepeleire J, Agossou C, Apers L, Zannou DM and Criel B. Lessons Learnt From the Experiences of Primary Care Physicians Facing COVID-19 in Benin: A Mixed-Methods Study. Front. Health Serv. 2022; 2:843058. 10.3389/frhs.2022.843058.

13. Von Pressentin KB, Mash RJ, Baldwin-Ragaven L, Botha RPG, Govender I, Steinberg WJ, et al. The influence of family physicians within the South African district health system: a cross-sectional study. Ann Fam Med. 2018 Jan;16(1):28–36. DOI: 10.1370/afm.2133.

14. Jenkins LS, Von Pressentin KB, Naidoo K, Schaefer R. The evolving role of family physicians during the coronavirus disease 2019 crisis: An appreciative reflection. Afr J Prm Health Care Fam Med. 2020;12:a2478. doi: 10.4102/phcfm.v12i1.2478

15. Codjia L, Jabot F, Dubois H. Évaluation du programme d’appui à la médicalisation des aires de santé rurales au Mali : accroître l’accès aux personnels de santé dans les zones rurales ou reculées : étude de cas N°2. Copenhague: OMS; 2010. French. https://apps.who.int/iris/handle/10665/44277.

16. Swanepoel M, Mash B, Naledi T. Assessment of the impact of family physicians in the district health system of the Western Cape, South Africa. Afr J Prim Health Care Fam Med. 2014 Dec;6(1):1–8. doi: 10.4102/phcfm.v6i1.695.

17. Von Pressentin KB, Mash RJ, Baldwin-Ragaven L, Botha RPG, Govender I, Steinberg WJ. The bird’s-eye perspective: How do district health managers experience the impact of family physicians within the South African district health system? A qualitative study. South African Fam Pract. 2018;60(1):13–20. doi: 10.1080/20786190.2017.1348047.

18. Flinkenflögel M, Sethlare V, Cubaka VK, Makasa M, Guyse A, De Maeseneer J. A scoping review on family medicine in sub-Saharan Africa: Practice, positioning and impact in African health care systems. Hum Resour Health. 2020;18(1):1–18. doi: 10.1186/s12960-020-0455-4.

19. Jan T, Aillet S. De Tananarive au Caire1: un aperçu du métier de médecin généraliste libéral dans le contexte des systèmes de santé locaux [Thèse de Médecine]. Bordeaux: Université Bordeaux 2 – Victor Segalen UFR des sciences médicales ; 2011. French.

20. Houenassi MD, Codjo LH, Dokoui D, Dohou SHM, Wanvoegbe A, Agbodande A, et al. Management of arterial hypertension in Cotonou city, Benin: general practitioners’ knowledge, attitudes and practice. Cardiovasc J Afr. 2016 Aug;27(4):e1–6. French. doi: 10.5830/CVJA-2015-094.

21. Iipinge SN, Pretorius L. The delivery and quality of sexually transmitted infections treatment by private general practitioners in Windhoek, Namibia. Glob J Health Sci. 2012 Aug;4(5):156–71. doi: 10.5539/gjhs.v4n5p156.

22. Beogo I, Darboe A, Adesanya OA, Rojas BM. Choosing between nurse-led and medical doctor-led from private for-profit versus non-for-profit health facilities: a household survey in urban Burkina Faso. PLoS One. 2018 Jul 1;13(7):e0200233. doi: 10.1371/journal.pone.0200233.

23. Mukiapini S, Bresick G, Sayed AR, Le Grange C. Baseline measures of primary health care team functioning and overall primary health care performance at Du Noon community health centre. Afr J Prim Health Care Fam Med. 2018 Sep;10(1):1–11. DOI: 10.4102/phcfm.v10i1.1458.

24. Bello K, De Lepeleire J, Agossou C, Zannou DM, Criel B. In-depth mapping of primary care physicians’ practices in four health districts in Benin: a mixed methods study [Preprint]. Research Square. Oct 2024. 10.21203/rs.3.rs-5217318/v1.

25. Palmer N, Mills A, Wadee H, Gilson L, Schneider H. A new face for private providers in developing countries: what implications for public health? Bull World Health Organ. 2003;81(4):292–7. DOI: 10.1016/j.socscimed.2003.12.015.

26. Wenghofer EF, Williams AP. Factors affecting physician performance: Implications for performance improvement and governance. Healthc Policy. 2009;5(2):e141–60. https://pmc.ncbi.nlm.nih.gov/articles/PMC2805145/.

27. Donabedian A. Evaluating the quality of medical care. Reprinted 1966 article. Milbank Q. 2005;83:691–729. 10.1111/j.1468-0009.2005.00397.x

28. Kringos, D.S., Boerma, W.G., Hutchinson, A. et al. The breadth of primary care: a systematic literature review of its core dimensions. BMC Health Serv Res. 2010; 10 (65). 10.1186/1472-6963-10-65.

29. Van Olmen J, Criel B, Bhojani U, Marchal B, Van Belle S, Chenge MF, et al. The health system dynamics framework: the introduction of an analytical model for health system analysis and its application to two case-studies. Health Cult Soc. 2012;2:1–21. doi: 10.5195/HCS.2012.71.

30. World Health Organization. The World Health Report 2008: Primary health care, now more than ever. Geneva: WHO press; 2008. https://apps.who.int/iris/handle/10665/43949

31. Word Health Organization, United Nations Children’s Fund. Primary health care measurement framework and indicators: monitoring health systems through a primary health care lens. Geneva: WHO press; 2022. https://iris.who.int/handle/10665/352201

32. Bresick G, Sayed AR, Le Grange C, Bhagwan S, Manga N. Adaptation and cross-cultural validation of the United States Primary Care Assessment Tool (expanded version) for use in South Africa. Afr J Prim Health Care Fam Med. 2015;7(1):783. doi: 10.4102/phcfm.v7i1.78.

33. Burt J, Abel G, Elmore N, Campbell J, Roland M, Benson J, et al. Assessing communication quality of consultations in primary care: initial reliability of the Global Consultation Rating Scale, based on the Calgary-Cambridge Guide to the Medical Interview. BMJ Open. 2014;4:e004339. doi: BMJ Open 2014;4:e004339. doi: 10.1136/bmjopen-2013-004339.

34. Lagarde M, Burn S, Lawin L, Bello K, Dossou J, Goufodji BS, et al. Exploring the Impact of Performance-Based Financing on Health Workers ’ Performance in Benin. Cotonou: World Bank; 2015.

35. Dedoose. [Accessed on October 30, 2024]. Available from: https://www.dedoose.com/

36. Braun V, Clarke V. Using thematic analysis in psychology. Qualitative Research in Psychology, 3(2), 77–101. 10.1191/1478088706qp063oa.

37. Dieleman M, Harnmeijer J. Improving health worker performance: in search of promising practices. Amsterdam: Royal Tropical Institute; 2006. https://www.kit.nl/wp-content/uploads/2018/08/1174_Improving-health-worker-performance_Dieleman_Harnmeijer.pdf

38. Mabuchi S, Sesan T, Bennett SC. Pathways to high and low performance: Factors differentiating primary care facilities under performance-based financing in Nigeria Health Policy Plan. 2018;33(1):41–58. doi: 10.1093/heapol/czx146.

39. Appiah-Denkyira E, Herbst CH, Soucat A, Lemiere C, Saleh K, eds. Towards interventions in human resources for health in Ghana: evidence for health workforce planning and result. Directions in Development. Washington, DC: World Bank; 2012. doi:10.1596/978-0-8213-9667-4.

40. Awases MH, Bezuidenhout MC, Roos JH. Factors affecting the performance of professional nurses in Namibia. Curationis. 2013;36(1):e1–8. doi: 10.4102/curationis.v36i1.108.

41. Irimu GW, Greene A, Gathara D, Kihara H, Maina C, Mbori-Ngacha D, et al. Factors influencing performance of health workers in the management of seriously sick children at a Kenyan tertiary hospital - Participatory action research. BMC Health Serv Res. 2014; 14(59). 10.1186/1472-6963-14-59.

42. Caza BB, Creary SJ. The construction of professional identity. In: Wilkinson A, Hislop D, Coupland C, editors. Perspectives on contemporary professional work: Challenges and experiences. Cheltenham: Edward Elgar Publishing; 2016. p. 259–85. doi: 10.4337/9781783475582.00022.

43. Van der Voort CTM, Van Kasteren G, Chege P, Dinant GJ. What challenges hamper Kenyan family physicians in pursuing their family medicine mandate? A qualitative study among family physicians and their colleagues. BMC Fam Pract. 2012;13(32):1–15 doi: 10.1186/1471-2296-13-32

44. Van Dormael M. L’introduction des sciences sociales dans une expérience de formation professionnelle des médecins de campagne au Mali. Colloque International Amades : Proceedings of the international colloquium of Anthropology and Medicine; 2007 Oct 25-27; Marseille, France. French. URL: http://www.strengtheninghealthsystems.be/doc/3/ref%203.6%20=%20ref%202.14%20introduction%20des%20sciences%20sociales%20-%20Mali.pdf

45. Mash R, Downing R, Moosa S, De Maeseneer J. Exploring the key principles of family medicine in sub-Saharan Africa: international Delphi consensus process. South African Fam Pract. 2008;50(3):60–5. doi: 10.1080/20786204.2008.10873720.

46. Cauli M. Contribution of humanities and social sciences to the training of healthcare professionals in Africa: An experience in the context of the mother and child Priority Solidarity Fund. Santé Publique. 2013;25:857–61. https://stm.cairn.info/journal-sante-publique-2013-6-page-857?lang=en.

47. Green LA, Yawn B, Lanier D, Dovey SM. The ecology of medical care revisited. N Engl J Med. 2001;344(26):2021–5. doi: 10.1056/NEJM200106283442611.

48. Moosa S. Family doctor leadership in African primary health care. Afr J Prim Health Care Fam Med. 2021; 13(1):1–2. doi: 10.4102/phcfm.v13i1.3198.

49. Kjeldmand D, Holmström I. Balint groups as a means to increase job satisfaction and prevent burnout among general practitioners. Ann Fam Med. 2008;6(2):138–45. doi: 10.1370/afm.813.

50. Travis P, Egger D, Davies P, Mechbal A. Towards better stewardship: concepts and critical issues. Geneva: World Health Organization; 2002. http://www.who.int/healthinfo/paper48.pdf

51. Moosa S, Peersman W, Derese A, Kidd M, Pettigrew LM, Howe A, et al. Emerging role of family medicine in South Africa. BMJ Glob Health. 2018;3:2–4. DOI: 10.1136/bmjgh-2018-000736.

52. Hellowell M, Myburgh A, Sjoblom M, Gurazada S, Clarke D. Covid-19 and the collapse of the private health sector: a threat to countries’ response efforts and the future of health systems strengthening? Geneva: Health Systems Governance Collaborative; 2020. https://hsgovcollab.org/en/blog/covid-19-and-collapse-private-health-sector-threat-countries-response-efforts-and-future53 Bregman P. The Right Way to Hold People Accountable. Harv Bus Rev 2016;:2–4.

53. Bregman P. The right way to hold people accountable. [Accessed on October 30, 2024]. Harv Bus Rev. 2016. Available from: https://hbr.org/2016/01/the-right-way-to-hold-people-accountable

54. Bitton A, Fifield J, Ratcliffe H, Karlage A, Wang H, Veillard JH, et al. Primary healthcare system performance in low-income and middle-income countries: a scoping review of the evidence from 2010 to 2017 BMJ Global Health 2019;4:e001551.

55. The World Bank. DataBank: World Development Indicators [Accessed on August 9, 2023]. Available from: https://databank.worldbank.org/reports.aspx?dsid=2&series=SP.URB.TOTL.IN.ZS

56. Nimpagaritse M, Korachais C, Meessen B. Effects in spite of tough constraints - A theory of change based investigation of contextual and implementation factors affecting the results of a performance based financing scheme extended to malnutrition in Burundi. PLoS One. 2020;15:e0226376. 10.1371/journal.pone.0226376.

57. 1000minds. Multi-Criteria Decision Analysis (MCDA/MCDM).. [Accessed on June 22, 2025]. Available from:https://www.1000minds.com/decision-making/what-is-mcdm-mcda#what-is-mcda.

