## Supplementary file 1 for "Factors supporting the performance of primary care physician practices in Benin: a multiple case study"

| **Variables** | **Set 1: Practice of quality improvement and public health activities** | **Set 2: Availability of essential services at affordable cost** | **Set 3: Satisfaction of the PCP** |
| --- | --- | --- | --- |
| Satisfaction of the PCP | *0.02* | *0.09* | **0.52** |
| Frequency to which non-clinical health activities are performed (staff supervision and training, quality improvement activities) | **0.68** | **0.42** | 0.11 |
| Health facility management activities (monitoring, reporting, etc.) | **0.63** | 0.28 | 0.16 |
| Cost of the PCPs’ consultation | *0.04* | **-0.62** | *0.02* |
| Number of patients consulted daily | -0.20 | 0.18 | 0.24 |
| Community-based outreach activities | 0.31 | 0.25 | 0.37 |
| Care for acute malnutrition | *0.04* | **0.46** | 0.38 |
| Practice of eutocic delivery | 0.16 | *0.00* | **0.45** |
| Orthopedic management of fractures | *0.02* | *-0.03* | *0.08* |
| Practice of health promotion activities | **0.48** | 0.24 | 0.30 |
| Practice of preventive clinical activities | *0.09* | 0.30 | 0.34 |
| Availability of essential services in the PCPs’ facilities | *0.05* | **0.41** | *0.00* |
| Preferred referral site for critical patients | -0.18 | *0.04* | *-0.04* |
| Completion of at least one continuing education course in the past 12 months | 0.26 | -0.11 | *-0.01* |

Supplementary file 1: Factor loading table for the three set of variables used to construct the criteria to approximatively appraise the PCPs performance
