## Supplementary file 2 for "Factors supporting the performance of primary care physician practices in Benin: a multiple case study"

Supplementary file 2: Detailed performance analysis for each case

### Introductory notes:

To analyse the performance of each case, the study team (principal investigator and research assistants) considered both the individual performance of the physician and the performance of the care team as a whole (using qualitative data and triangulating viewpoints of all stakeholders).

**In terms of individual performance, the team considered the following dimensions:**

- Technical performance (as observed by the research team and as perceived by stakeholders). For the observation the team chose the following thresholds to assess the physician's performance (based on medians and quartiles):
  - Percentage of recommended clinical procedures below 75%: low score
  - Percentage of recommended clinical procedures between 75% and 85%: average score
  - Percentage of recommended clinical procedures above 85%: good score
- Performance in patient-centred care (as observed by the research team and as perceived by stakeholders). For observation purposes, the team chose the following thresholds to assess the physician's performance (based on medians and quartiles)
  - Overall communication score below 15/20: low score
  - Overall communication score between 15/20 and 17/20: average score
  - Overall communication score above 17/20: good score
- The physician's contribution to the performance of the healthcare team and the health centre

**In terms of overall performance, the team considered the following parameters**

- Technical performance, taking into account both the physician's performance and the performance perceived by stakeholders
- Performance for patient-centred care, considering both the performance of the physician and the performance of the team as perceived by stakeholders
- The accessibility (financial, geographical, etc.) of the services offered by the case
- Continuity of service
- The relationship between the physician and the care team and the community
- The case's contribution to strengthening the local health system

Table 1 summarizes the results of this analysis. Details foreach case are available in the following pages.

Table 1: Summary table of performance analysis for the 8 cases

|  | Case 1 | Case 2 | Case 3 | Case 4 | Case 5 | Case 6 | Case 7 | Case 8 |
| --- | --- | --- | --- | --- | --- | --- | --- | --- |
| Technical effectiveness | Average | Poor | Good | Average | Good | Average | Average | Average |
| Patient-centeredness (PCP) | Average | Average | Average | Good | Good | Good | Good | Average |
| Accessibility | Average | Average | Average | Average | Average | Average | Good | Poor |
| Continuity | Average | Average | Average | Average | Good | Good | Good | Average |
| Community engagement | Average | Poor | Good | Good | Average | Good | Good | Good |
| PCP's capacity to strengthen the primary care team and the health centre | Poor | Poor | Average | Good | Good | Good | Good | Average |
| Practice's contribution to strengthen the local health system | Average | Average | Good | Good | Average | Good | Good | Good |

### **Case 1**

Case 1 focuses on a physician with no academic qualifications beyond a doctorate in medicine (general practitioner). She had 7 years' professional experience at the time of collection. She did not indicate that she was enrolled in a postgraduate training course. The health facility where this physician practises and which was included in case 1 is a private (faith-based) health centre located in an urban area with a population of around 57,962. The health centre employs 2 physicians, 7 nurses, 0 midwives and 5 orderlies. The physician in case 1 does not manage the health centre.

#### Performance summary

Overall, the **technical efficiency performance of case 1 was average**. The physician in case 1 had an average individual performance in terms of technical efficiency, with a percentage of recommended clinical procedures performed of 76%. From the stakeholders' point of view, his performance in patient management was average. According to the people interviewed, its technical efficiency is improving (compared to when it started at the centre). The technical performance of its health facility was judged to be good by stakeholders, and many patients travel miles to come to the centre for treatment.

**In terms of patient-centred care, case 1's overall performance was average**. On an individual level, the physician in case 1 performed poorly, with an overall communication score of 14.7/20 at observation. However, the stakeholders felt that she gave a good welcome and listened well to patients, and the reception at the centre was judged to be good.

**In terms of accessibility, the performance of Case 1 was average.** The services provided in the health centre (including medical consultation fees) were considered affordable by all the stakeholders interviewed. In addition, the two physicians are permanently available during the centre's opening hours (working days from 8am to 4pm). However, local people complained that they could not access care at the centre at night or at weekends. The long queues to see the physician are another barrier to access noted by local people.

**For continuity of care, the performance of case 1 is average.** The physician gives follow-up appointments to monitor the patient's state of health, particularly for patients with chronic conditions. However, there is no information system that enables patients' records to be kept efficiently over the long term (system based on care diaries and paper registers). This leads to gaps in the follow-up of patients' appointments, as we were able to see for ourselves during our observations, with two diabetic patients who had gaps in their follow-up of several months and two years respectively.

**In terms of community relations, the performance of case 1 is average.** Because of the nature of the centre (denominational), there is a certain link with the community through religious activities (e.g. morning prayers to which patients and carers are invited). In addition, the centre has a fairly loyal patient flow. However, we did not note any activities carried out in the community by the centre or the physician, nor a system for maintaining regular contact with the local authorities.

**In terms of the physician's contribution to the performance of the care team and the health facility, performance is poor.** According to the stakeholders, the case 1 physician's leadership skills are average. Stakeholders reported poor availability of medicines and limited equipment at the centre. However, we cannot link these factors to the physician's performance because she is not directly involved in managing the centre. In addition, the physician's contribution to strengthening the care teams and to quality assurance is weak. The physician is not very involved in activities to improve the quality of care. She sometimes consults the paramedics and there is good collaboration, but we did not note any real involvement on the part of the physician in strengthening the team's capacities. Existing capacity-building activities consist of training courses or meetings on patient management. However, these are not very regular.

**In terms of the case's contribution to strengthening the local health system, performance was average.** We noted good liaison with the district hospital when a case was referred, thanks in particular to referral forms. Collaboration between the health centre and the health zone management team is going well. However, this collaboration seems to be mainly administrative, with little real discussion to build a shared vision. The physician herself does not seem to be personally involved in these exchanges with the zone management team. In terms of achieving the results targeted by the local health system, it is difficult for us to assess the case's contribution. The centre does not have its own geographical area. However, the centre is very busy, receiving around thirty patients a day. The members of the community interviewed appreciated the fact that they had easy access to a physician and even said that they came specifically to this centre because they could find a physician at an affordable cost. This centre therefore seems to meet a need for quality, affordable primary care, as patients travel long distances for treatment.

### **Case 2**

Case 2 focuses on a physician with no academic qualifications beyond a doctorate in medicine (general practitioner). She had just over 2 years' professional experience at the time of data collection. She did not indicate that she was enrolled in a postgraduate training course. The health facility where this physician practises and which was included in case 2 is a public health centre located in an urban area of around 57,691 inhabitants. The health centre employs 2 physicians, 9 nurses, 7 midwives and 5 orderlies. The physician studied in case 2 does not manage the health centre. Furthermore, unlike the other physicians in our sample working in a public health facility, the physician in case 2 is not a civil servant but rather a service provider who is paid by the centre according to the time spent in the health facility.

#### Performance summary

Overall, **the technical efficiency performance of case 2 was poor**. The physician in case 1 had a poor individual performance in terms of technical efficiency, with a percentage of recommended clinical procedures performed of 73%. From the point of view of the stakeholders, his performance in patient management was average, with some appreciating his performance positively and others being more reserved. The technical performance of the health facility included in case 2 was judged to be average by the stakeholders.

**In terms of patient-centred care, the overall performance of case 2 was average**. On an individual level, the physician in case 1 had an average performance, with an overall communication score of 17/20 on observation. Stakeholders felt that she provided a good welcome and listened well to patients. However, the reception at the centre was judged to be poor.

**In terms of accessibility, the performance of Case 2 was average.** The services provided in the health centre were considered relatively expensive by the stakeholders interviewed. They acknowledged that the costs were lower than in some private facilities in the locality, but they were still expensive for their means and more expensive than other public health centres in the locality. Patients also complained of long queues. However, on the positive side, stakeholders felt that the physician in case 2 was helping to reduce the cost of treatment (unlike centres without a physician) by making better prescriptions. In addition, there is a system of on-call physicians so that there is always a physician available in the centre.

**For continuity of care, the performance of case 1 is average.** The physician gives follow-up appointments to monitor the patient's state of health. But there is no continuity over time, and no long-term relationship with patients, because the on-call teams change. So a patient who has been given an appointment may come back and find another physician or another team. What's more, we didn't see any effective information system to facilitate patient follow-up.

**In terms of community relations, the performance of case 2 is weak.** The centre (including the physician) does not carry out any activities in the community. The community members interviewed complained several times about this lack of involvement in the community. They would have liked the centre to organise awareness-raising activities and for the centre's staff to listen more carefully to the needs of the community.

**In terms of the physician's contribution to the performance of the care team and the health facility, the performance of case 2 is poor.** According to the stakeholders, the case 2 physician's ability to lead is average. Stakeholders reported limited equipment and infrastructure at the centre. Most medicines are bought outside the centre. However, we cannot link these factors to the physician's performance, as she is not at all involved in managing the centre. In terms of strengthening the care teams and quality assurance, the physician in case 2 was not very involved in activities to improve the quality of care. Collaboration with the paramedics, while not strained, was rather distant, with each simply doing his or her job. Activities to improve the quality of care (which are not very regular and mainly involve training and meetings on patient management) are managed by the centre's administrative authorities, with no real role for the physician in question or his colleague.

**In terms of coordination with other levels of the healthcare system, case 2 performed averagely.** We noted good liaison with the district hospital when a case was referred, thanks in particular to referral forms. Collaboration between the health centre and the health zone management team was good. However, this collaboration seems to be mainly administrative. The physician herself is not personally involved in these exchanges with the zone management team. Here again, it is difficult for us to assess the case's contribution to achieving the results targeted by the local health system. The centre does not have its own dedicated geographical area.

### Case 3

Case 3 focuses on a physician with a specialist diploma in a clinical speciality (specialist) but with a general practice. He had 10 years' professional experience at the time of data collection. The health facility where this physician practises and which was included in case 3 is a public health centre located in a semi-urban area with a population of around 14,844. The staff providing clinical care (including the physician studied) in this health centre comprise 1 physician, 6 nurses, 2 midwives and 6 orderlies. The physician studied in case 3 manages this health centre. He is a civil servant.

#### Performance summary

**Case 3's technical performance was judged to be good**. The physician in case 3 performed well individually in terms of technical efficiency. When the consultation was observed, he performed 86% of the recommended clinical procedures, and most of the stakeholders interviewed were positive about his efficiency. Stakeholders also judged that the technical quality of care offered by the health centre involved in case 3 was good.

**Case 3's performance in terms of patient-centred care was average**. The physician's individual performance (perceived and observed) was good, with an overall communication score of 18.9/20 and the majority of participants believing that he was good at welcoming patients and listening to them. At the health centre, however, people complained about the poor reception they received (especially when the physician was away).

**In terms of accessibility, the performance of case 3 is average**. On the one hand, the physician helps to reduce costs for patients by improving diagnosis and limiting prescriptions to the minimum necessary. In addition, affordability at his centre was judged to be good. The cost of a general medical consultation at this centre is 500 FCFA (not including other services). However, the physician in case 3 is not available all the time (at night, at weekends, and at various times during the week), due to the fact that he works in several health facilities at the same time and the fact that he does not live in the locality of his centre. He is the only physician at the centre.

**In terms of continuity of care, the performance was average**, because apart from making appointments for the follow-up of patients he sees in consultation, we did not note any other aspects of continuity of care. Also, the DPH felt that once a patient had been referred to another level of care, the patient was no longer his responsibility. As far as the information system is concerned, patients' clinical information is compiled on paper media (care diaries or registers), which makes it difficult to retain information and monitor patients.

**In terms of community relations, Case 3 performed well.** The physician is good at listening to the needs of the community and has very good relations with it. He listens to people's complaints and takes action accordingly, sends his staff to listen to the community's grievances, often consults the local authorities, and works with government representatives from other sectors. The centre also has a management committee made up of community representatives. This committee is supposed to support the centre in managing its finances and ensuring that the needs of the population are taken into account in the services offered and in the way services are organised. The centre also relies on community relays.

**In terms of the physician's contribution to the performance of the health care team and the health facility, the performance of Case 3 was average.** On the positive side, several stakeholders felt that the physician (who is in charge of the health centre) has a good ability to lead his teams and the health centre. He has also managed to mobilise resources for his centre from various stakeholders. He organises activities to improve the quality of care, such as meetings at which the centre's performance is discussed with recommendations for improvement, and certain topics are presented to improve the knowledge of health workers. The centre's staff testified that the physician helps them to access various training courses organised by the health zone and that he supervises and coaches them in their work. The staff say they have learned a lot from him. However, we were able to observe that these activities to improve the quality of care did not seem to have any real influence on the reception at the centre. What's more, although the staff report that there is good co-operation between themselves and with the physician, the relationship between the physician and his staff is more of a "command and control" type, with occasional tensions. In terms of medicines, equipment and infrastructure, the local population reported poor availability of medicines and inadequate availability of equipment and infrastructure. However, these last two aspects seem to be linked to the general context of public health establishments, rather than to the specific performance of the physician or centre.

**In terms of the case's contribution to strengthening the local health system, Case 3 performed well**. There is good liaison with the district hospital when a case is referred, thanks to referral forms and telephone calls. This liaison is even easier with the physician himself, who works at the district hospital (where most cases are referred) and is a specialist. Collaboration with the health zone management team goes very well, as the physician himself is a member of this team. As a result, he helps to define the health objectives for the zone and ensures that his activities are in line with these objectives. In addition, the case's contribution to achieving the results targeted by the local health system is clearly visible because the health centre has a well-defined geographical area and population of responsibility. We had several testimonies that the physician had helped to increase attendance at the health centre and the coverage of health interventions.

### **Case 4**

Case 4 is centred on a physician who had no academic qualifications after the doctorate in medicine (general practitioner). He had 4 years' professional experience at the time of collection. However, he had been a nurse for a few years before starting medical school. He did not indicate that he was enrolled in a postgraduate training course. The health facility where this physician practises and which was included in case 4 is a public health centre located in a semi-urban area with a population of around 10,527. The health centre employs 1 physician, 1 nurse, 1 midwife and 4 orderlies. The physician studied in case 4 manages this health centre. He is a civil servant.

#### Performance summary

**In terms of technical efficiency, case 4 had an average technical performance**. The physician's individual performance was good, with an 88% completion rate for recommended clinical procedures. Stakeholders also felt that he was effective in managing patients. However, according to the stakeholders interviewed, the care provided in the health facility is of average technical efficiency.

**In terms of patient-centred care, Case 4 performed well**. The physician in case 4 had a good overall communication score of 17.63/20. Stakeholders felt that this physician was good at welcoming patients and listening to them. At the level of the centre, the welcome given to patients was judged to be good.

**In terms of accessibility, the performance of Case 4 is average**. The centre was very affordable. In addition, the physician helps to reduce costs according to the patients because he prescribes effective products from the outset, which avoids prolonging the illness and buying several prescriptions. However, the physician is not available all the time, especially when he is busy with other activities at the zone office or elsewhere. He is the only physician at the centre and is not replaced during his absence. However, he can be contacted by telephone

**In terms of continuity of care, the performance of case 4 was average**. The physician tries to maintain a certain continuity of care through follow-up appointments. However, he was unable to keep track of all his patients, given his many responsibilities. In addition, detailed information on patients is only compiled in care diaries or, at best, in paper files, which does not make it easy to keep track of these patients. Apart from the health information system, where statistics are compiled, we have not identified any digitised information system for patient records.

**In terms of community relations, case 4's performance is good**, with a good ability to listen to the needs of the community, including consultation with the local authorities and collaboration with government representatives from other sectors. According to the community members interviewed, the physician in case 4 is very available to listen and give advice to the community. He is also involved in community development activities such as setting up a sports group or a forum where community leaders can discuss local problems. The centre also relies on community relays and has a management committee.

**In terms of the physician's contribution to the performance of the care team and the health facility, the performance of case 4 is good**. According to the people interviewed, the physician has a good ability to manage the centre. He is also able to mobilise resources from the community. However, the centre's equipment and infrastructure were deemed inadequate by the stakeholders, and there were periods of disruption in the availability of medicines. However, these two problems are common to all public health facilities. To strengthen the care teams and quality assurance, the physician often organises and takes part in activities to improve the quality of care, such as meetings at which the centre's performance is discussed with the paramedics. He also supervises his staff. In addition, the population has testified that since their arrival at the centre, the performance of the paramedical health workers and that of the centre in general has improved.

**Case 4 performed well in terms of its contribution to strengthening the local health system**. The physician and his health centre liaise well with the district hospital when a case is referred, thanks in particular to referral forms. Collaboration with the health zone management team is going very well, as the physician himself is a member of this team. As a result, he helps to define the health objectives for the zone and is responsible for ensuring that his activities are in line with these objectives. In terms of contributing to achieving the results targeted by the local health system, the centre has a well-defined geographical area. We have had testimonies showing an increase in attendance at the centre and in the coverage of interventions since his arrival at the centre.

### **Case 5**

Case 5 focuses on a physician with no academic qualifications beyond a doctorate in medicine (general practitioner). He had 8 years' professional experience as a physician at the time of collection. However, he had been a laboratory technician for a few years before starting medical studies. He is currently training for a medical speciality. The health facility where this physician practises and which was included in case 5 is a private clinic located in an urban area of about 69,799 inhabitants. The staff providing clinical care (including the physician studied) in this health centre include 2 physicians, 3 nurses, 2 midwives and 3 orderlies. The physician in case 5 runs this private clinic.

#### Performance summary

**Case 5's technical performance was judged to be good**. The physician in case 5 performed well individually in terms of technical efficiency, with a percentage of recommended clinical procedures performed of 85%. Most of the stakeholders interviewed were also positive about his efficiency. The technical efficiency of the health facility in case 5 was also judged to be good.

**In terms of patient-centred care, the performance of case 5 was good.** The physician's individual performance (perceived and observed) was good, with an overall communication score of 19.0/20 and the majority of participants believing that he was a good host and listener to patients. The public also judged that patients were well received and well listened to at the health facility in case 5.

**In terms of accessibility, the performance of case 5 is average**. The physician in case 5 is very available when patients ask for him and there is an on-call system so that there is always a physician in the centre. In addition, the physician and his colleagues are available for home consultations. The care team reported that they try to apply "social rates" for people who cannot afford to pay for all the care (for example, they may not include the cost of the physician's travel when they do home consultations). However, members of the community felt that care at the health facility was expensive and that you needed to have money or health insurance to access it. The cost of a general medical consultation at this centre is between 2,000 and 5,000 CFA francs (not including other services).

**Case 5 performed well in terms of continuity of care**. The physician systematically schedules follow-up appointments for patients, and there is a system for following up and reminding patients of their appointments, especially those with serious or chronic conditions. There is a digital system for keeping patient records, facilitating continuity over time.

**In terms of community relations, the case's performance was average**. The physician maintains good relations with the local authorities. He also organises a number of awareness-raising and screening activities for illnesses (especially chronic illnesses) in the community and provides home consultations. However, we did not notice any real consultation with the community about their health problems. What's more, the way the centre is organised (no community management committee, private company) and the cost of services limit Case 5's ability to respond to the community's needs.

**In terms of the physician's contribution to the performance of the care team and the health facility, the performance of case 5 is good.** According to the people interviewed, the physician has good leadership skills. He created the centre in association with colleagues. He is able to mobilise resources (most of them private) to develop the centre and is also very active in publicising the centre. Equipment and medicines are also readily available. In terms of strengthening the care teams and quality assurance, the Case 5 physician often organises staff meetings to discuss the cases received and the quality of care. The physician is also personally involved in monitoring and supervising the care provided by paramedics and some of his medical colleagues. According to the physician himself, these activities to improve the quality of care are essential to the centre's survival. According to the other staff at the centre, the physician's involvement in the quality of care had a positive impact on their own performance. However, they were less appreciative of the fact that his rigour was perhaps a little too great at times. Here too, the relationship between the physician and the paramedics is one of "command and control".

**In terms of its contribution to strengthening the local healthcare system, Case 5 performed moderately well**. We noted good liaison with the referral hospital, with referral forms drawn up for referrals. However, these referrals are not often made to the district hospital and there is no strong collaboration between the centre and the district hospital (or even the hospital to which cases are often referred). However, the physician does have contacts with various specialists, which he activates when necessary. The physician also has good personal contacts with the local health authorities and is active in medical associations. However, we did not note any systematic collaboration with the zone management team in defining and achieving health objectives. In terms of Case 5's contribution to achieving the results targeted by the local health system, the centre contributes to screening for chronic diseases, which was cited as an unmet need by several health authorities we met in the locality. But it was difficult for us to really assess the contribution of the case because the centre shares its geographical area with several other public and private health centres. Finally, at the time of data collection, the centre was barely two years old. The physician and the other co-founders of the centre aspired to turn it into a polyclinic with several specialities and sophisticated laboratory and radiology analyses. This raises questions about the centre's affordability and the consistency of its activities with the health zone's future development plan.

### **Case 6**

Case 6 focuses on a physician who, after completing his doctorate in medicine, obtained a university diploma in general community medicine (community general practitioner). He had 14 years' professional experience at the time of the survey. He is currently training for a specialty in public health. The health facility where this physician practises is a private community medical centre located in a rural area with a population of around 22,291. The staff providing clinical care (including the physician studied) in this health centre include 1 physician (2 at certain times), 2 nurses, 0 midwives and 4 orderlies.

The physician in case 6 runs this centre.

#### Performance summary

**Case 6's technical performance was judged to be average.** Case 6 had an average individual performance in terms of technical efficiency, with an 81% completion rate for recommended clinical procedures. However, all the stakeholders interviewed felt that he was effective in managing patients. The care provided in the health facility concerned by case 6 was also judged to be effective by all the stakeholders interviewed.

**In terms of patient-centred care, Case 3 performed well**. The physician's overall consultation score was 17.7/20. Stakeholders felt that patients were well received and listened to by both the physician and the health centre.

**In terms of accessibility, the performance of case 6 was average**. According to the participants, the centre has good financial accessibility and the physician helps to reduce the cost of care by diagnosing illnesses early, for example. The cost of a general medical consultation at this centre is between 1,500 and 2,000 CFA francs (not including other services). However, the physician is the only physician in the centre and is not available on site at night, at weekends, or when he is busy with other activities. He can be contacted by telephone and is replaced when he is absent for long periods.

**In terms of continuity of care, Case 6 performed well**. The physician has been at the centre for several years now and keeps track of the patients and families in his care over time. He actively follows up patients' appointments, especially chronic cases. The health centre has an information system for monitoring patients with chronic conditions, although this system is not digitalised. The centre looks for lost patients, relying on community contacts and the good knowledge of families by the physician and his team.

**In terms of community relations, Case 6 performed well.** All the stakeholders reported a good ability to listen to the needs of the community. This is achieved both by the physician and by the other staff through consultations with the local authorities, discussions during awareness-raising sessions and community visits, and the monitoring of ailments that frequently appear in the community. The centre also relies on community relays and has a management committee. It has to be said that the local authorities were very involved in the creation of the centre, and we noted that they were very proud of it. Finally, the physician and the health centre contribute to the development of the locality and the community, for example by helping to draw up birth certificates for newborn babies, and by making the health centre's borehole available to the local population.

**In terms of the physician's contribution to the performance of the health care team and the health facility, the performance of case 6 is good.** The physician in case 6 set up the centre (with the support of the French NGO Santé Sud and the local communities). Stakeholders felt that he had a good capacity to run the centre. Although there are still shortcomings, the physician has gradually improved the centre's technical facilities and infrastructure over the years. There is also a good availability of medicines at the centre. In terms of strengthening the care teams and quality assurance, the physician regularly organises training or refresher sessions for his staff on common ailments, especially during epidemics or seasonal upsurges of endemic diseases (such as malaria). Staff also reported that he supervises and coaches them when they are working. They felt that he "knows how to show others how to do things". We also noted that despite the clear hierarchy between the physician and his staff, there was a good relationship between them and a team spirit.

**In terms of the case's contribution to strengthening the local health system, Case 6 performed well**. The physician and his staff ensure good liaison with the referral hospital (which is usually the district hospital) by means of referral forms and calls to the hospital before referring. In addition, the people testified that in the event of a referral, the physician personally ensures that the patient is treated at the hospital and follows up by calling the hospital's carers and visiting the patient at the hospital. The physician in case 6 has a good relationship with the district management team. He is invited to take part in activities organised by this team and helps to define the health objectives for the zone. As for the case's contribution to achieving the results targeted by the local health system, the health centre has a well-defined geographical area and population of responsibility. According to the information gathered, the presence of the centre in the locality has filled a geographical gap in the availability of health facilities in the area. The centre has also provided the population with an alternative to health facilities and to people offering (poor quality) care illegally. The centre has helped to increase the utilisation rate and coverage of interventions in the area, to reduce referrals (due to better first-line care and a reduction in the severity of cases presenting for treatment), to combat epidemics, and so on.

### **Case 7**

Case 7 is centred around a physician who, after completing her doctorate in medicine, obtained a university diploma in community general medicine (community general practitioner). She had 12 years' professional experience at the time of collection. She did not indicate that she was enrolled in a postgraduate training course. The health centre where this physician practises is a private community medical centre located in a rural area (and isolated due to the poor state of the access roads) of around 15,804 inhabitants. The health centre employs 1 physician, 1 nurse, 0 midwife and 4 orderlies. The physician in case 7 is the head of the centre.

#### Performance summary

**Case 7's technical performance was judged to be average.** Physician case 7 had an average individual performance in terms of technical efficiency, with a percentage of completion of recommended clinical procedures at 80%. However, the stakeholders interviewed felt that she was effective in managing patients. The care provided in the health facility concerned by case 6 was also judged to be effective by the stakeholders interviewed.

**In terms of patient-centred care, Case 7 performed well.** The physician performed well, with an overall observation communication score of 19.28/20. Stakeholders at the centre also reported that patients were well received and well listened to (regardless of socio-economic level or physical condition).

**In terms of accessibility, Case 7 performed well**. We noted that the physician was always available because she lived in the centre. When she is away for several days, she is replaced by another physician. What's more, there is good financial accessibility, adapted to the financial means of the population covered by the centre.

**In terms of continuity of care, the performance of Case 7 is good**, because there is long-term monitoring of patients and their families. The physician makes regular home visits to check on the condition of patients with serious or chronic conditions, or who are elderly. The centre also has a digital information system where patient information is recorded and appointments are reminded, particularly for chronic cases.

**In terms of community relations, Case 7 performed well.** The physician has a good ability to listen to the needs of the community. She is in constant contact and often discusses health issues with members of the community, including local leaders and managers from other sectors (e.g. teachers). It regularly organises awareness campaigns and community visits (carried out by itself or its staff). It contributes to the development of the locality and the community by, for example, making the centre's water borehole available to the community, organising activities for young people, raising awareness of the damage caused by pesticides, etc.

**In terms of the physician's contribution to the performance of the health care team and the health facility, the performance of case 7 is good**. The physician in case 7 set up the centre (with the support of the French NGO Santé Sud and the local communities). We noted a good capacity to manage the centre, with ongoing work to improve the technical facilities within the limits of the centre's resources. The centre also has a good supply of medicines. In addition, the physician often organises training and refresher courses for paramedical staff, especially at the start of an epidemic or a season when there is an upsurge in cases of endemic diseases such as malaria. Staff and patients reported that she had taught her staff the basics of how to properly receive patients and provide care. We also noted good relations between the physician and paramedics and a team spirit, even a community spirit.

**Case 7 performed well in terms of its contribution to strengthening the local health system.** The physician liaises well with the referral hospital (which is usually the district hospital) through referral forms and calls to the hospital before referring. People also said that the physician often visited his referred patients at the hospital and that having a referral form from the physician made it easier for them to be treated at the hospital (unlike if they went there on their own). The physician in case 7 also has good relations with the district management team and contributes to the activities organised by this team (vaccination, epidemiological surveillance, etc.). We also noted good relations with the specialists and good coordination with them in the management of chronic cases. Case 7 also contributes to achieving the results targeted by the local health system. The community medical centre has a well-defined geographical area and population of responsibility. The centre has filled a geographical gap in the availability of health facilities in the health zone. The centre has also enabled the population to be offered an alternative to health facilities and to people offering (poor quality) care illegally. Other positive results include improved coverage of interventions in the area, especially for isolated populations, a reduction in home births and obstetric emergencies, and a reduction in the severity of cases presenting for care.

### **Case 8**

#### Case 8 is centred around a physician with no academic qualifications beyond a doctorate in medicine (general practitioner). She had 10 years' professional experience at the time of collection. She did not indicate that she was enrolled in a postgraduate training course. The health facility where this physician practises and which was included in case 8 is a public health centre located in a semi-urban area of around 15,927 inhabitants. The health centre employs 1 physician, 4 nurses, 1 midwife and 7 orderlies. The physician studied in case 8 runs this health centre. She is a civil servant.

#### Performance summary

Overall, the **technical efficiency performance of case 8 was average**. The physician in case 8 had a poor individual performance in terms of technical efficiency, with a percentage of recommended clinical procedures performed of 68%. From the stakeholders' point of view, her performance in patient management was average, with some believing she was effective and others expressing reservations. Stakeholders also rated the technical efficiency of the care provided at the facility as average.

**In terms of patient-centred care, Case 8's performance was average**. On an individual level, the physician in case 8 performed averagely, with an overall communication score of 17/20 in the observation. However, the stakeholders felt that she gave a good welcome and listened well to patients. At centre level, the welcome given to patients was judged to be good.

**In terms of accessibility, Case 8 performed poorly.** Stakeholders appreciated the fact that the physician is open to receive patients and can often be reached by telephone. In addition, she lives close to the centre, which means she can drop in and see patients outside normal working hours. However, she is not available all the time because of her administrative responsibilities and the various activities that this responsibility requires of her. Also, people at the centre complained about the long queues before being seen and the high cost of care at the centre (in relation to their financial means).

**For continuity of care, the performance of case 8 is average.** The physician gives appointments to the patients she sees. But she does not always manage to keep these appointments, due to lack of availability. However, she tries to organise follow-up for patients with chronic conditions by arranging for her patients to be seen regularly by members of her team, when she is unable to do so herself. Finally, the information system for compiling patient information is based on care diaries and paper registers, which does not facilitate continuity of care.

**In terms of relations with the community, case 8's performance is good, with a good ability to listen to the needs of the community,** including good consultation with the local authorities and collaboration with government representatives from other sectors (through frequent meetings and exchanges). The Case 8 physician also often works with community leaders to combat epidemics, carry out other public health activities and advise and raise awareness among the population. The centre also relies on community relays and has a management committee.

**In terms of the case physician's contribution to the performance of the care team and the health facility, Case 8's performance is average.** According to the stakeholders, the case 8 physician's leadership skills are average. The population appreciated the fact that the centre was clean and well organised. However, the centre's equipment and infrastructure were deemed inadequate by the stakeholders, and there were periods of disruption in the availability of medicines. These two problems are common to all public health facilities. To strengthen the care teams and quality assurance, the physician often organises and gets involved in activities to improve the quality of care, such as meetings at which the centre's performance is discussed with the paramedics. She also supervises her staff and pays close attention to the application of standard treatment procedures. However, the centre's technical performance was rated as average. In addition, relations between the physician and paramedics were average, and the physician was criticised for being too attached to standards.

**In terms of its contribution to strengthening the local healthcare system, case 8 performed well**. We have noted good liaison with the district hospital when a case is referred, thanks in particular to referral forms. Collaboration with the health zone management team is going very well, as the physician herself is a member of this team. As a result, she helps to define the health objectives for the zone and is responsible for ensuring that her activities are in line with these objectives. We also noted good coordination with specialist physicians in the management of chronic pathologies. The centre has a well-defined geographical area in terms of its contribution to achieving the results targeted by the local health system. We heard evidence that the physician is helping to improve health indicators.
