## Supplementary file 3 for "Factors supporting the performance of primary care physician practices in Benin: a multiple case study"

Supplementary file 3: Cross case analysis matrix

| **Observed factors** | **Case 1** | **Case 2** | **Case 3** | **Case 4** | **Case 5** | **Case 6** | **Case 7** | **Case 8** |
| --- | --- | --- | --- | --- | --- | --- | --- | --- |
| **Values** | - Commitment to serve the community - Professionalism - Altruism (and commitment to help people) | - Commitment to serve the community - Professionalism - Altruism as official value but environment incentivizes to make profit | - Commitment to serve the community - Professionalism - Altruism as official value but job description incentivizes to make profit - Respect for standards and hierarchy. | - Commitment to serve the community - Professionalism - Altruism as official value but job description incentivizes to make profit | - Commitment to serve the community - Professionalism - Profit-oriented values - respect for standards and hierarchy. | - Commitment to serve the community - Professionalism - Altruism (and commitment to help people) | - Commitment to serve the community - Professionalism - Altruism (and commitment to help people) | - Commitment to serve the community - Professionalism - Altruism as official value but job description incentivizes to make profit - Respect for standards and hierarchy. |
| **Preparation of the PCP to practice at the first- line** | - Little preparation to practice at the first-line | - Little preparation to practice at the first-line | - Good preparation for community work and implementation of the national health policy. - Extensive technical training | - Good preparation for community work and implementation of the national health policy. | - Good preparation for technical quality and facility management | - Very good preparation in many aspects | - Very good preparation in many aspects | - Good preparation for community work and implementation of the national health policy. |
| **Continuing education** | - No systematic continuing education program | - No systematic continuing education program | - No systematic continuing education program - Received a training session the past 12 months, but not systematically geared towards an identified need | - No systematic continuing education program - Received a training session the past 12 months, but not systematically geared towards an identified need | - No systematic continuing education program - Received a training session the past 12 months, but not systematically geared towards an identified need | - No systematic continuing education program - Received a training session the past 12 months, but not systematically geared towards an identified need | - No systematic continuing education program | - No systematic continuing education program - Received a training session the past 12 months, but not systematically geared towards an identified need |
| **Equipment and infrastructure** | - Insufficiency in equipment and. infrastructure | - Insufficiency in equipment and. infrastructure | - Insufficiency in equipment and. infrastructure - Managed to work around the insufficiency | - Insufficiency in equipment and. infrastructure | - Good equipment and. infrastructure | - Insufficiency in equipment and. infrastructure - Managed to work around the insufficiency | - Insufficiency in equipment and. infrastructure | - Insufficiency in equipment and. infrastructure |
| **Support from the hierarchy, peers or associations** | - Support provided by the head of the health center and other senior staff - Support consists in feedback and advice regarding her performance, leading to a progressive improvement in the PCP’s performance | - Little support provided to the PCP or the rest of the primary care team | - Support provided by the district health management teams, especially the district medical officer - Support includes connecting the PCPs with community leaders, explaining their roles to the PCPs, providing tips for the facility management, and access to various policy documents - Received coaching from the district medical officer | - Support provided by the district health management teams, especially the district medical officer - Support includes connecting the PCPs with community leaders, explaining their roles to the PCPs, providing tips for the facility management, and access to various policy documents | - Some support from peers and certain health authorities - But lack of financial support | - Support from peers and MGCs’ association - Support consists in regular meetings and other platforms to share advice and opportunities, financial support to set up the facility, and various collaborations. - Initial support from the French NGO “Santé Sud” | - Peers and MGCs’ association - Support consists in regular meetings and other platforms to share advice and opportunities, financial support to set up the facility, and various collaborations. - Initial support from the French NGO “Santé Sud” | - Support provided by the district health management teams, especially the district medical officer - Support includes connecting the PCPs with community leaders, explaining their roles to the PCPs, providing tips for the facility management, and access to various policy documents |
| **Leadership mandate given to the PCP and the degree of autonomy allocated to exert it** | - No leadership mandate leading to few effort to strengthen the primary care team or facility | - No leadership mandate leading to few effort to strengthen the primary care team or facility | - Official mandate to lead the health facility and coordinate health activities in the commune - A certain degree of autonomy but limited by the obligation to respond to their own hierarchy and by the little room of manoeuvre for holding the primary care team accountable | - Official mandate to lead the health facility and coordinate health activities in the commune - A certain degree of autonomy but limited by the obligation to respond to their own hierarchy and by the little room of manoeuvre for holding the primary care team accountable | - Owner of the health facility, thus a de facto mandate to lead it - Great autonomy for taking decisions | - Owner of the health facility, thus a de facto mandate to lead it - Great autonomy for taking decisions | - Owner of the health facility, thus a de facto mandate to lead it - Great autonomy for taking decisions | - Official mandate to lead the health facility and coordinate health activities in the commune - A certain degree of autonomy but limited by the obligation to respond to their own hierarchy and by the little room of manoeuvre for holding the primary care team accountable |
| **Modalities of the financing of the PCPs’ practices** | - Some financing from the church but a great part is financed by the income generated by the health services - Still some pressure for financial survival leading to effort to seek patient’s satisfaction - Services costs are kept acceptable thanks to the church’s support but also costs containment strategies (no permanent opening) | - Partially financed by the State and partially by the income generated by the health services - Less financial pressure than the other cases (large part f the staff is financed by the State) | - Partially financed by the State and partially by the income generated by the health services - Less financial pressure than the other cases (large part f the staff is financed by the State) | - Partially financed by the State and partially by the income generated by the health services - Less financial pressure than the other cases (large part f the staff is financed by the State) | - Entirely financed by private funds, mainly income generate by the services provided - High pressure for financial survival, leading to actively seek patient’s satisfaction but also to increased cost | - Some financing from the NGO “Santé Sud” but mainly at the beginning, a great part is financed by the income generated by the health services - Still some pressure for financial survival leading to effort to seek patient’s satisfaction but also and increase in costs (even if much less than case 5) - Services costs are kept acceptable thanks to costs containment strategies (use of generic drugs, only one physician, etc.) and an improved financial management with the help of a professional accountant | - Some financing from the NGO “Santé Sud” but mainly at the beginning, a great part is financed by the income generated by the health services - Still some pressure for financial survival leading to effort to seek patient’s satisfaction but also and increase in costs (even if much less than case 5) - Services costs are kept acceptable thanks to costs containment strategies (use of generic drugs, only one physician, etc.) and an improved financial management with the help of a professional | - Partially financed by the State and partially by the income generated by the health services - Less financial pressure than the other cases (large part f the staff is financed by the State)s |
| **Fulfilment of the PCP's personal needs** | - Good. - Has all evenings and afternoons free. - Is well integrated in her workplace community but not very much in the community the practice covers. - Want to embrace other professional activities (or have a specialty training) | - Average - Needs to do many shifts. - Not well integrated neither in her workplace community nor on the broader community the practice covers - Want to embrace other professional activities (or have a specialty training) | - Average. - Very busy schedule - Integrated in the community is working in but not very much in a personal way. - Lives in an adjacent city at about 1 hour from his workplace (difficult for his family to leave in the town of his workplace) | - Very good - Very busy schedule - Lives (with his family) and is well-integrated in the community he is working in (for instance has a regular sport schedule with community members, coaches young people, etc.) | - Good - Very busy schedule - Lives (with his family) near the clinic and has some community level activities, - but told us that he doesn’t have much time for himself | - Good. - Well integrated in the community he is working in and participates in many community activities.   However, no longer lives in the community but in an adjacent city at about 1 hour from his workplace (difficult for his family to leave in the town of his workplace) | - Very good - Lives and well-integrated in the community she is working in (many leisure activities with the community members, coaches young people, etc.) - Manages to have time for personal life activities | - Good - Lives and is well integrated in the community she is working in. and participates in many community activities. - Recently had her family joining her and she said that this is a factor that helped her to stay in the community and be more focused on her work |
| **Regulation** | - Not accessed as the regulation is the same for all the PCPs in the country | - Not accessed as the regulation is the same for all the PCPs in the country | - Not accessed as the regulation is the same for all the PCPs in the country | - Not accessed as the regulation is the same for all the PCPs in the country | - Not accessed as the regulation is the same for all the PCPs in the country | - Not accessed as the regulation is the same for all the PCPs in the country | - Not accessed as the regulation is the same for all the PCPs in the country | - Not accessed as the regulation is the same for all the PCPs in the country |
| **Accountability mechanisms** | - Some accountabilities towards the health authorities, but limited to administrative issues - No specific population or area of responsibility - Little pressure to achieve key health systems’ performance indicators - PCPs’ role not clearly stated, apart from the general understanding that the physicians have from tacit knowledge | - Some accountabilities towards the health authorities, but limited to administrative issues - No specific population or area of responsibility - Little pressure to achieve key health systems’ performance indicators. - PCPs’ role not clearly stated, apart from the general understanding that the physicians have from tacit knowledge | - Clear accountability mechanisms from the PCPs to the health authorities, including clear job description and deliverables expected, clear geographic area of responsibility. - Most deliverables expected from the case and the PCP are geared towards and support facility management, community orientation and attainment of the health systems’ performance indicators. - Some deliverables are geared toward quality of care for the patients, but measurement mechanisms fail to assess them | - Clear accountability mechanisms from the PCPs to the health authorities, including clear job description and deliverables expected, clear geographic area of responsibility. - Most deliverables expected from the case and the PCP are geared towards and support facility management, community orientation and attainment of the health systems’ performance indicators. - Some deliverables are geared toward quality of care for the patients, but measurement mechanisms fail to assess them | - Some accountabilities towards the health authorities, but limited to administrative issues - No specific population or area of responsibility - Little pressure to achieve key health systems’ performance indicators - PCPs’ role not clearly stated, apart from the general understanding that the physicians have from tacit knowledge - Some accountability towards the patients because those attending this health facility often chose to do so and have the means to choose other centers. | - Clear accountability towards health authorities, peers, and community, thanks to a MGC charter signed by the PCPs, the municipal authorities and the health authorities. - MGC charter states the roles and the deliverables expected from the PCPs and the case 6 has a clear geographic area of responsibility. - A strong accountability towards the community with community leaders providing regular feedback to the PCP - Accountability mechanisms help the case to monitor its performance and improve it | - Clear accountability towards health authorities, peers, and community, thanks to a MGC charter signed by the PCPs, the municipal authorities and the health authorities. - MGC charter states the roles and the deliverables expected from the PCPs and the case 7 has a clear geographic area of responsibility. - Some accountability towards the community with community leaders providing regular feedback to the PCP - Accountability mechanisms help the case to monitor its performance and improve it | - Clear accountability mechanisms from the PCPs to the health authorities, including clear job description and deliverables expected, clear geographic area of responsibility - Most deliverables expected from the case and the PCP are geared towards and support facility management, community orientation and attainment of the health systems’ performance indicators. - Some deliverables are geared toward quality of care for the patients, but measurement mechanisms fail to assess them |
| **Collaboration with community leaders and other public officials** | - Weak collaboration with community leaders and other key stakeholders | - Weak collaboration with community leaders and other key stakeholders | - Actively maintain a good collaboration with municipal authorities and other key stakeholders - Help the PCPs to mobilize resources and to facilitate community activities. - Participation to multisectoral dialogue platforms helping the PCPs to effectively contributes to strengthen the local health system and to mutualize resources | - Actively maintain a good collaboration with municipal authorities and other key stakeholders - Help the PCPs to mobilize resources and to facilitate community activities. - Participation to multisectoral dialogue platforms helping the PCPs to effectively contributes to strengthen the local health system and to mutualize resources | - Some collaboration with community leaders and other key stakeholders, but not very strong | - Actively maintain a good collaboration with municipal authorities and other key stakeholders - Help the PCPs to mobilize resources and to facilitate community activities. - Participation to multisectoral dialogue platforms helping the PCPs to effectively contributes to strengthen the local health system and to mutualize resources | - Actively maintain a good collaboration with municipal authorities and other key stakeholders - Help the PCPs to mobilize resources and to facilitate community activities. - Participation to multisectoral dialogue platforms helping the PCPs to effectively contributes to strengthen the local health system and to mutualize resources | - Actively maintain a good collaboration with municipal authorities and other key stakeholders - Help the PCPs to mobilize resources and to facilitate community activities. - Participation to multisectoral dialogue platforms helping the PCPs to effectively contributes to strengthen the local health system and to mutualize resources |
| **Relationship with the rest of the primary care team and leadership style of the PCPs** | - Tacitly seen as the leader of the primary care team (even if no mandate) - Mere cordial relationship - PCP and other cadres work in silo, which hampers knowledge sharing - The curative consultations are only provided by the PCPs Both the PCPs and the health facility are only available 8h a day and 5 days in a week. | - Tacitly seen as the leader of the primary care team (even if no mandate) - PCP and other cadres work in silo, which hampers knowledge sharing - The curative consultations are only provided by the PCPs - The continuity of the services is ensured by a duty system where the there is a PCP in the facility 24h/7 - The permanent presence of physician increases the cost for the facility and potentially for the patients | - Officially seen as the leader of the primary care team - Hierarchical relationship between the PCP and the primary care team - Command and control leadership style from the PCP, encouraging the staff to comply to the PCPs’ instructions but only in his presence - Nurse- - practitioners share responsibility for curative consultations with the PCP - The nurse-practitioners refer to the PCPs patients with condition two complex for them to handle but not needing a referral to the district hospital. - This task sharing may negatively impact the quality of the care provided to patient as in this case the PCP’s capacity to strengthen the rest of the primary care team is average | - Officially seen as the leader of the primary care team - Hierarchical relationship between the PCP and the primary care team, but still some flexibility in this hierarchy - Nurse-practitioners share responsibility for curative consultations with the PCP - This compensates for the PCPs’ limited availability for patient care at a lower cost than if the PCPs’ absence was compensated by another physician. - The nurse-practitioners refer to the PCPs patients with condition two complex for them to handle but not needing a referral to the district hospital | - Officially seen as the leader of the primary care team - Hierarchical relationship between the PCP and the primary care team - Command and control leadership style from the PCP encouraging the staff to comply to the PCPs’ instructions and provide good quality care - The curative consultations are only provided by the PCPs - The continuity of the services is ensured by a duty system where the there is a PCP in the facility 24h/7 - The permanent presence of physician increases the cost for the facility and potentially for the patients | - Officially seen as the leader of the primary care team - Strong collaboration between the PCPs and the rest of the primary care team. - Person-centered approach in the leadership style encouraging knowledge sharing, teamwork, and shared values - The curative consultations and other clinical acts such as deliveries are primarily the responsibility of the PCP. - However, nurse-practitioners also provide curative consultations during short absences of the PCP, with the PCP calling for another colleague only for prolonged absence. - This ensures the continuity of the services - This task sharing has a little impact on the quality of the care provided to patient as the primary care team is average care is provided by a PCP most of the time and the PCPs has a good capacity to strengthen his team | - Officially seen as the leader of the primary care team - Strong collaboration between the PCPs and the rest of the primary care team. - Person-centered approach in the leadership style encouraging knowledge sharing, teamwork, and shared values - The curative consultations and other clinical acts such as deliveries are primarily the responsibility of the PCP. - However, nurse-practitioners also provide curative consultations during short absences of the PCP, with the PCP calling for another colleague only for prolonged absence. - This ensures the continuity of the services - This task sharing has a little impact on the quality of the care provided to patient as the primary care team is average care is provided by a PCP most of the time and the PCPs has a good capacity to strengthen her team | - Officially seen as the leader of the primary care team - Hierarchical relationship between the PCP and the primary care team - Command and control leadership style from the PCP encouraging the staff to stick to the standards set by the Ministry of health, but not innovating - Nurse-practitioners share responsibility for curative consultations with the PCP - The nurse-practitioners refer to the PCPs patients with condition two complex for them to handle but not needing a referral to the district hospital. - This task sharing may negatively impact the quality of the care provided to patient as in this case the PCPs’ capacity to strengthen the rest of the primary care team is average |
| **Workload and availability of the PCP** | - Average number of patients per day: 10 - No significant workload in terms of non-clinical duties. | - Average number of patients per day: 8 - No significant workload in terms of non-clinical duties. | - Average number of patients per day: 9 - A significant workload for non-clinical (including administrative and managerial) duties, leading to long working hours - Limited availability of the PCP due to the non-clinical duties compensated by the nurse-practitioners compensate for the PCPs’ limited availability for patient care | - Average number of patients per day: 7 - A significant workload for non-clinical (including administrative and managerial) duties, leading to long working hours - Limited availability of the PCP due to several non-clinical (including administrative) duties - The nurse-practitioners compensate for the PCPs’ limited availability for patient care | - Average number of patients per day: 7 - A fairly significant workload for non-clinical (including administrative and managerial) duties, leading to long working hours - PCP still available for patient care but also supported by other PCPs | - Average number of patients per day: 7 - Some workload for non-clinical duties but not very high - Part of the administrative duty is taken over by a professional administrator | - Average number of patients per day: 4 - Some workload for non-clinical duties but not very high | - Average number of patients per day: 7 - A significant workload for non-clinical (including administrative and managerial) duties, leading to long working hours - Limited availability of the PCP due to the non-clinical duties compensated by the nurse-practitioners compensate for the PCPs’ limited availability for patient care |
| **Contextual aspect** | - Urban area - Competition with other centers but not very strong because of the affordable prices - Private facility owned by the church - Community served is middle-income to poor people - Health insurance not common among patients | - Urban area - Competition with other centers - Public facility but with a particular status - Community served is middle-income to poor people - Health insurance not common among patients | - Urban area - Public facility - Moderate competition with other centers - Community served is middle-income to poor people (but more poor than middle income) - Health insurance almost inexistant among patients | - Semi-urban area - Public facility - Moderate competition with other centers - Community served is middle-income to poor people (but more poor than middle income) - Health insurance almost inexistant among patients | - Urban area - High competition with other centers - Private facility owned by a small group of individuals (including the PCP). - Community served is middle-income to rich people - Health insurance very common among patients | - Rural area - Little competition with other centers (inexistant if we only consider practices with physicians) - Private facility owned by the PCP - Community served is poor people - Health insurance almost inexistant among patients | - Rural area - Little competition with other centers (inexistant if we only consider practices with physicians) - Private facility owned by the PCP - Community served is poor people - Health insurance almost inexistant among patients | - Semi-urban area - Public facility - Moderate competition with other centers - Community served is middle-income to poor people (but more poor than middle income) - Health insurance almost inexistant among patients |
